# Understanding COVID-19 testing behaviour in England through a sociodemographic lens

**DOI:** 10.1101/2023.10.26.23297608

**Authors:** Sumali Bajaj, Siyu Chen, Richard Creswell, Reshania Naidoo, Joseph L.-H. Tsui, Olumide Kolade, George Nicholson, Brieuc Lehmann, James A Hay, Moritz U. G. Kraemer, Ricardo Aguas, Christl A. Donnelly, Tom Fowler, Susan Hopkins, Liberty Cantrell, Prabin Dahal, Lisa J. White, Kasia Stepniewska, Merryn Voysey, Ben Lambert, the EY-Oxford Health Analytics Consortium

**Author notes:** Contributed equally (Sumali Bajaj, Siyu Chen, Richard Creswell, Reshania Naidoo, Joseph L.-H. Tsui). The EY-Oxford Health Analytics Consortium membership list is at the end of the article.

## Abstract

**Background:** Understanding underlying mechanisms of heterogeneity in test-seeking and reporting behaviour can help to protect the vulnerable and guide equity-driven interventions. Using COVID-19 testing data for England and data from community prevalence surveillance surveys (REACT-1 and ONS-CIS) from October 2020 to March 2022, we investigated the relationship between sociodemographic factors and testing behaviours in England.

**Methods:** We used mass testing data for lateral flow device (LFD; data for 290 million tests performed and reported) and polymerase chain reaction (PCR) (data for 107 million tests performed and returned from the laboratory) tests made available for the general public, provided by date, self-reported age and ethnicity at lower tier local authority (LTLA) level. Using a mechanistic causal model to debias the PCR testing data, we obtained estimates of weekly SARS-CoV-2 prevalence by self-reported ethnic groups and age groups for LTLAs in England. This approach to debiasing the PCR (or LFD) testing data also estimated a testing bias parameter defined as the odds of testing in infected versus not infected individuals, which would be close to zero if the likelihood of test seeking (or seeking and reporting) was the same regardless of infection status. Using confirmatory PCR data, we estimated false positivity rates, sensitivity, specificity, and the rate of decline in detection probability by PCR by sociodemographic groups. We also estimated the daily incidence allowing us to determine the fraction of cases captured by the testing programme.

**Findings:** From March 2021 onwards, individuals in the most deprived regions reported approximately half as many LFD tests per-capita than those in the least deprived areas (Median ratio [Inter quartile range, IQR]: 0·50 [0·44, 0·54]). During October 2020 – June 2021, PCR testing patterns were in the opposite direction (Median ratio [IQR]: 1·8 [1·7, 1·9]). Infection prevalences in Asian or Asian British communities were considerably higher than those of other ethnic groups during the Alpha and Omicron BA.1 waves. Our estimates indicate that the England COVID-19 testing program detected 26% - 40% of all cases (including asymptomatic cases) over the study period with no consistent differences by deprivation levels or ethnic groups.

PCR testing biases were generally higher than for LFDs, which was in line with the general policy of symptomatic and asymptomatic use of these tests. During the invasion phases of the Delta and Omicron variants of concern, the PCR testing bias in the most deprived populations was roughly double (ratio: 2·2 and 2·7 respectively) that in the least. We also determined that ethnic minorities and older individuals were less likely to use confirmatory PCR tests through most of the pandemic and that there was possibly a longer delay in reporting a positive LFD test in the Black populations.

**Interpretation:** Differences in testing behaviours across sociodemographic groups may be reflective of the relatively higher costs of self-isolation to vulnerable populations, differences in test accessibility, digital literacy, and differing perception about the utility of tests and risks posed by infection. Our work shows how mass testing data can be used in conjunction with surveillance surveys to identify gaps in the uptake of public health interventions at fine scale levels and by sociodemographic groups. It provides a framework for monitoring local interventions and yields valuable lessons for policy makers in ensuring an equitable response to future pandemics.

**Funding:** UK Health Security Agency.

## Introduction

More unequal societies have poorer health outcomes^1^. The COVID-19 pandemic likely further exacerbated the impact of existing societal inequalities, yet our understanding of the impact of the pandemic across different parts of society remains poor. Understanding the underlying causes of heterogeneity in infection risk and disease burden is essential to protect the vulnerable and to guide equity-driven interventions in real-time.

In England, testing for SARS-CoV-2 played an important role during the COVID-19 pandemic, allowing the government to monitor disease transmission and forming the cornerstone of self-isolation policies. However, it is unclear whether testing was delivered and taken up equitably across society^2–6^. The UK government rolled-out mass testing in response to the COVID-19 pandemic, with the initial testing response starting in March 2020^7^. The testing programme component of NHS Test and Trace (NHSTT) played an integral role through its various testing services. Since May 2020, PCR tests were available for testing swabs from suspected COVID-19 cases; LFD tests for COVID-19 were rapidly developed and evaluated for use between Dec 2020 and April 2021.^8,9^. LFD testing was first rolled-out to key populations, for example, those working in healthcare, and subsequently to the wider population through a universal testing offer. The universal testing offer was a major component of the national testing programme, with a key aim to expand asymptomatic LFD testing and was delivered via various channels, with rollout to the general public from 9 April 2021^10^.

We had unprecedented access to COVID-19 testing data for England as part of a UK Health Security Agency (UKHSA) commissioned retrospective evaluation of England’s testing programme over an evaluation period of October 2020 – March 2022^9^. Here, we present analyses of these data which quantify biases in test uptake and reporting patterns across different sociodemographic groups, including those based on age, self-reported ethnic group and a measure of deprivation. We also estimate SARS-CoV-2 infection prevalence over time and space for sociodemographic groups, which additionally allows us to estimate the reach of the national testing programme. Our analysis focusses only on the Pillar 2 testing data, which includes PCR and LFD tests made available to the general public (not including tests made available to individuals with clinical need, and health and care workers through Pillar 1).

## Methods

### Data

The testing data were provided by UKHSA in the context of a retrospective evaluation of the delivery of England’s COVID-19 testing programme and its impact on public health outcomes^11^. Our analysis period covered 1^st^ October 2020 - 30^th^ March 2022. LFD data relied on self-reporting, meaning that our analysis focused on self-reported results. In contrast, the PCR data are results from registered and submitted samples of individuals. These Pillar 2 data were provided by date by a range of categories (including self-reported age and ethnicity) and at lower tier local authority (LTLA) level. Individuals self-reported their ethnicities according to five groupings: “Asian or Asian British” (abbreviated throughout as “Asian ethnic groups” or “Asian people”), “Black; African; Black British or Caribbean” (abbreviated as “Black ethnic groups” or “Black people”), “White”, “Mixed or Multiple ethnic groups”, and “Other ethnic group”; the Office for National Statistics (ONS) also publishes population estimates for these groups, which we used. We present results for the first three ethnic groups in the main section since the population sizes for these were considerably higher; in the supplementary materials, we report results for the remaining categories. We used publicly available data on population size estimates by LTLAs, LTLA and ethnic groups, and age groups, as well as deprivation indices/levels.

We used weekly LTLA-level and weekly age group specific community surveillance random sampling survey data from the REal-time Assessment of Community Transmission (REACT-1) study which performed PCR tests of cross-sectional random samples of individuals approximately every month from 1^st^ May 2020 to 31^st^ March 2022^12^. We also used publicly available national weekly prevalence estimates from the ONS COVID-19 Infection Survey (CIS) which repeatedly tested a sample of people^13^. ONS prevalence estimates were used only to estimate national incidence (see Methods below).

### Variant periods

The following were the time periods during which the Alpha, Delta and Omicron variants of concern (VOCs) were dominant in England i.e. when the relative frequency of each VOC exceeded 90% nationally (see Figure 1A, Figure S1):

- Alpha: 30^th^ January 2021 to 24^th^ April 2021
- Delta: 5^th^ June 2021 to 4^th^ December 2021
- Omicron: 1^st^ January 2022 to 14^th^ May 2022 (but we ended our Omicron period in March 2022 since this was the end of our study period)

**Figure 1:**
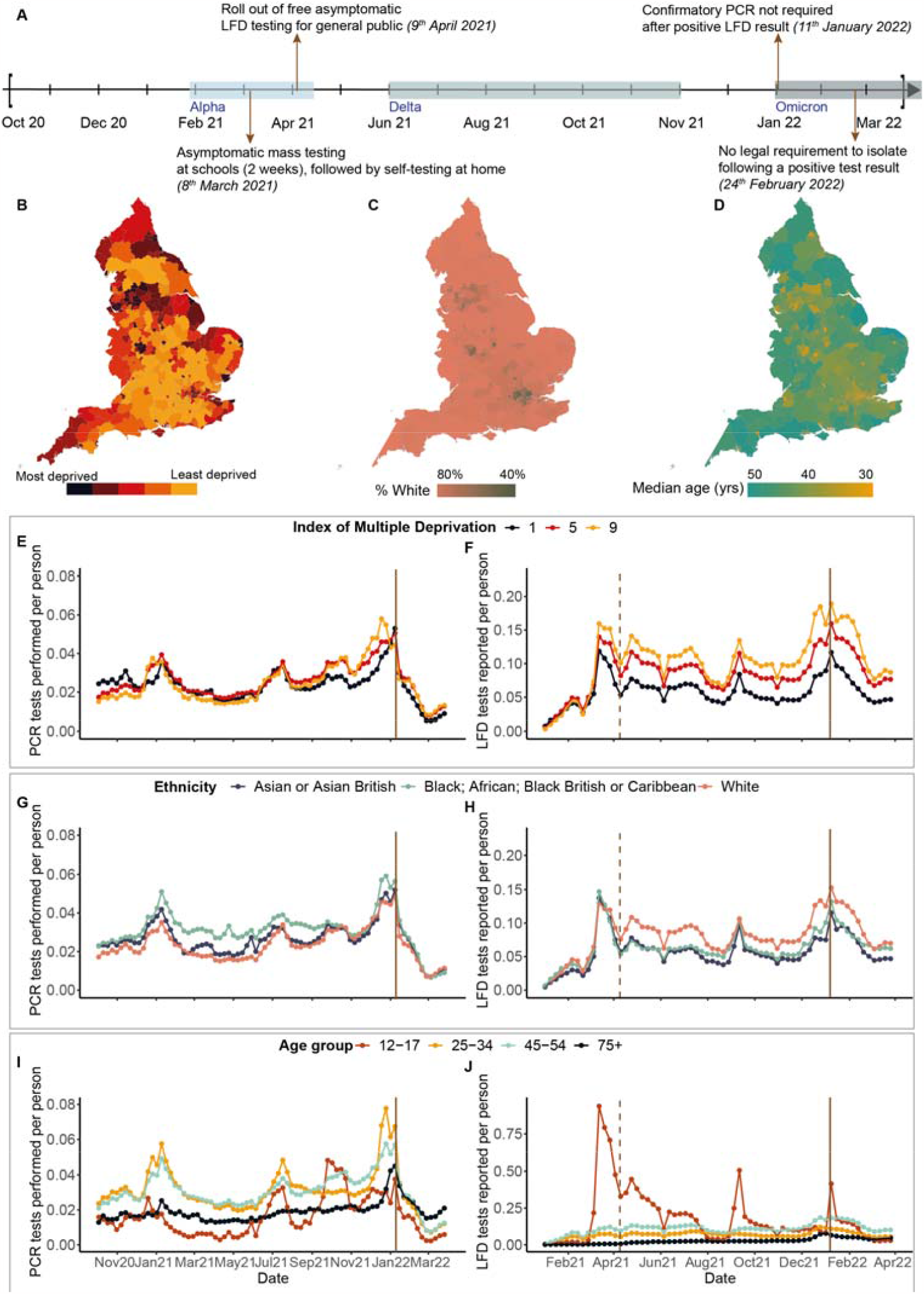
(A) Timeline of the Alpha, Delta, and Omicron variants of concern and other important dates mentioned in our study (B) Spatial distribution of deprivation across England according to the rank of population weighted average of scores for the LSOAs in a given LTLA. (C) Proportion of White people across LTLAs. (D) Median age across LTLAs of England. (E, F) Weekly number of PCR, LFD tests per person by deprivation levels. All LTLAs were assigned deprivation levels from 1 to 9. (G, H) Weekly number of PCR, LFD tests per person by three ethnic groups (see Figure S10 for the other two ethnic groups). (I, J) Weekly number of PCR, LFD tests per person by four age groups (see Figure S9 for the other age groups). The solid vertical line corresponds to the date (11^th^ January 2022) after which confirmatory PCR tests were no longer required following a positive LFD test, and the vertical dashed line corresponds to the date (9^th^ April 2021) when free asymptomatic testing was made available to all individuals in England. See Figure S11 for variation in tests per person by LTLAs and sociodemographic groups.

### Prevalence estimates and testing bias

Community surveillance random sampling survey testing programmes such as REACT-1 and ONS CIS provide a gold-standard measure of infection prevalence; they have large sample sizes nationally and can produce precise estimates of prevalence at this level. At combinations of fine geographic or other scales (LTLA level and sociodemographic groups), the effective sample size is much reduced, particularly because prevalence was generally low (<10%). Conversely, at finer scales, there is a wealth of Pillar 2 PCR and LFD tests conducted, but the naïve prevalence estimates derived from these tests may be biased. This is because PCR tests were used for symptomatic testing or as part of test-to-release protocols^14^, and even though LFD tests were recommended for asymptomatic testing, they likely suffer from reporting bias (unlike PCR). We implement and extend the debiasing methodology of Nicholson et al.^15^ which in addition makes use of large volumes of Pillar 2 PCR and LFD tests. This method uses swab results from the REACT-1 study to correct for biases in mass PCR testing data and thus produces fine scale PCR swab positivity rate estimates at a higher precision than is possible just from the community surveillance survey. For ease of readability, we use we refer to PCR swab positivity rate as prevalence. The method estimates testing bias at a coarse scale where there are high numbers of both community surveillance survey tests and mass tests. These biases are then propagated to the nested finer spatial scales to estimate prevalence, making use of the sufficiently large volume but biased mass testing data at fine scales. The testing biases estimated represent the log odds of testing in infected versus not infected individuals. For example, if the probability of performing a PCR test was the same regardless of whether people were infected, then the biases would equal zero (= log (1)). So, a large bias corresponds to a higher chance of an infected individual performing a PCR test compared with an uninfected individual, which would result in overestimated prevalence using the biased testing data. We also estimate the LFD test reporting bias using data on reported LFD tests at the coarse and fine scales. We term this as bias in context of using these data naively for surveillance purposes; however, we recognise that the testing programme was primarily a public health intervention rather than a surveillance programme and did not aim to use PCR or LFD testing data to determine SARS-CoV-2 prevalence.

Using the debiasing method described above, we generate two novel sets of prevalence estimates for each LTLA: one set, stratified by ethnic groups (Figure S2); another, by age group (Figure S3). We also use the method to estimate the LTLA specific prevalence (Figure S4) and testing bias for deprivation levels (Index of Multiple Deprivation IMD).

### False positivity and test specificity and sensitivity

We collated a subset of our Pillar 2 testing data comprising all reported LFD positive results which were followed up with a PCR test taken within three days of the reported positive. These data allowed us to estimate the false positive rate for LFD tests, assuming PCR tests were a gold standard. Using the total number of people reporting LFD positives, we can additionally determine the probability that individuals reporting an LFD positive result subsequently took a PCR test, as was generally mandated throughout the period under observation, with two exceptions: from January to March 2021 and after January 2022^16^ when confirmatory PCR testing was temporarily suspended given the high COVID-19 prevalence meaning LFD tests likely had low false discovery rates. Using our estimates of prevalence and false positivity rates, we estimated LFD test specificity and sensitivity across different age and ethnic groups throughout the pandemic.

### Modelling the decay in PCR detection rate after reporting an LFD positive

Using the data described above, we modelled how the probability of detecting SARS-CoV-2 infection by PCR after reporting a positive LFD result declined according to the time elapsing between reporting the LFD test and subsequent testing by PCR. We determined this decay rate for the different ethnic groups for monthly time windows throughout the study period.

### Estimating SARS-CoV-2 incidence and the case detection ratio from prevalence time series

An uptick in cases may signal future increases in hospitalisations and deaths. However, case data are imperfect, being influenced by changes in both the true incidence of infection and changes in testing / test reporting behaviour. We estimated daily incidence in a Bayesian paradigm, assuming the following probabilistic relationship between infections and prevalence:

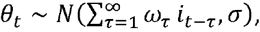

where *θ*_*t*_denotes prevalence on day *t* and *i*_*t*_is the incidence on the same day; 0 ≤ *ω*_*τ*_ ≤ 1 is the probability of being PCR positive given an infection which began *τ* days ago and *N*(*μ, σ*) represents a normal distribution with mean *μ* and standard deviation *σ*.

We assume that within 15-day rolling blocks, the reported PCR and LFD positive counts for the Pillar 2 mass testing data at time *t* were a to-be-estimated constant fraction of the true infection incidence, given by the following relationship:

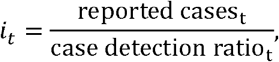

with case detection ratio_*t*_ corresponding to the 15-day block that time *t* corresponds to. We describe our priors in supplementary materials.

The availability of two prevalence datasets (the REACT-1 surveys and the UK ONS at the national level only) made it possible for us to infer the true incidence of infection in England and to compare this with the reported cases derived from the national mass testing programme. We used the LTLA-specific prevalences estimated using Pillar 2 PCR data and REACT-1 data as described above to estimate true incidence at the LTLA level and then aggregated it to get true national incidence estimates. Using the ONS prevalence estimates for England, we estimated another set of national incidences. We report time series of the average national case detection ratios derived from REACT-1 and ONS-CIS datasets defined as the fraction of PCR and LFD positive tests in the Pillar 2 dataset compared to the estimated true incidence, by LTLA-specific sociodemographic groups.

Details of all methods are in the Supplementary Materials.

## Results

### Trends in testing uptake and reporting

Over our study period, a total of 290 million LFD tests were reported with 2% of these results positive; and 107 million PCR tests were registered with 13% of tests positive (Figure S5). Of the 7 million positive LFD tests reported, 2.4 million were confirmed by a PCR test with 68% of these on the same day (Table 1 and Figure S6), and 89% returning a positive result.

**Table 1:** Summary of Pillar 2 mass testing data for England between 1^st^ October 2020 and 31^st^ March 2022. * includes tests where there were data with information on LTLAs or ethnicity or age.

|  | Number of tests | Number of positive results (%) |
| --- | --- | --- |
| PCR tests performed* |  |  |
| With LTLA | 106,986,026 | 14,021,052 (13·1) |
| With ethnicity | 106,831,682 | 13,933,106 (13·0) |
| With age | 103,681,935 | 13,554,671 (13·1) |
| LFD tests reported* |  |  |
| With LTLA | 290,462,387 | 7,007,391 (2·4) |
| With ethnicity | 290,227,237 | 6,991,160 (2·4) |
| With age | 286,525,766 | 6,441,966 (2·2) |
| **Confirmatory PCR tests** | **Number of LFD tests** | **Number of PCR positive results (%)** |
| LFD positives followed by PCR | 2,328,414 | 2,069,391 (88·9) |
| Within same day | 1,598,213 | 1,416,229 (88·6) |
| After one day | 513,567 | 461,159 (89·8) |
| After two days | 209,143 | 185,511 (88·7) |
| After three days | 7491 | 6492 (86·7) |

From October 2020 – June 2021, individuals in the most deprived areas (IMD=1) (Figure 1B) performed almost double (Median ratio [Inter quartile range, IQR]: 1·8 [1·7, 1·9]) the number of PCR tests per capita compared to those in the least deprived (IMD=9) (Figure 1E), and we speculate that this is due to the higher prevalence in deprived areas (Figure S7A) during this period. Linear regression models for the PCR tests per capita suggest that differences in prevalence may explain some of the variation in testing across deprivation levels (Figure S7B, C). But myriad other factors likely also influenced testing, including individuals’ age, their occupation, their risk perception and their vaccination status, and a more comprehensive approach would be necessary to probe further the observed differences in testing levels^17^. LFD test reporting displayed the opposite and more pronounced trend, where those in the most deprived areas reported the fewest LFD tests per capita, which held even after the roll-out of free asymptomatic testing on 9^th^ April 2021 (Figure 1A). From March 2021, the median number of LFD tests reported per capita in the most deprived areas was roughly half that in the least deprived (Median ratio [IQR]: 0·50 [0·44, 0·54]) despite more deprived areas having the highest prevalence [Figure S7A] – a finding qualitatively similar to the results of the Liverpool COVID-19 study^18^. This may be partly explained by the disproportionately low digital access and resources to report LFDs, as well as lack of adequate support to isolate if testing positive. Those in Black ethnic groups had generally higher PCR testing coverages throughout the analysis period (Figure 1G). Since our prevalence estimates do not indicate a consistently higher prevalence in Black ethnic groups (Figure 2A), it is possible that this reflects a greater risk of symptomatic disease, access to tests through workplace, and could also be partly explained by the ramping up of targeted community testing^19,20^.

**Figure 2:**
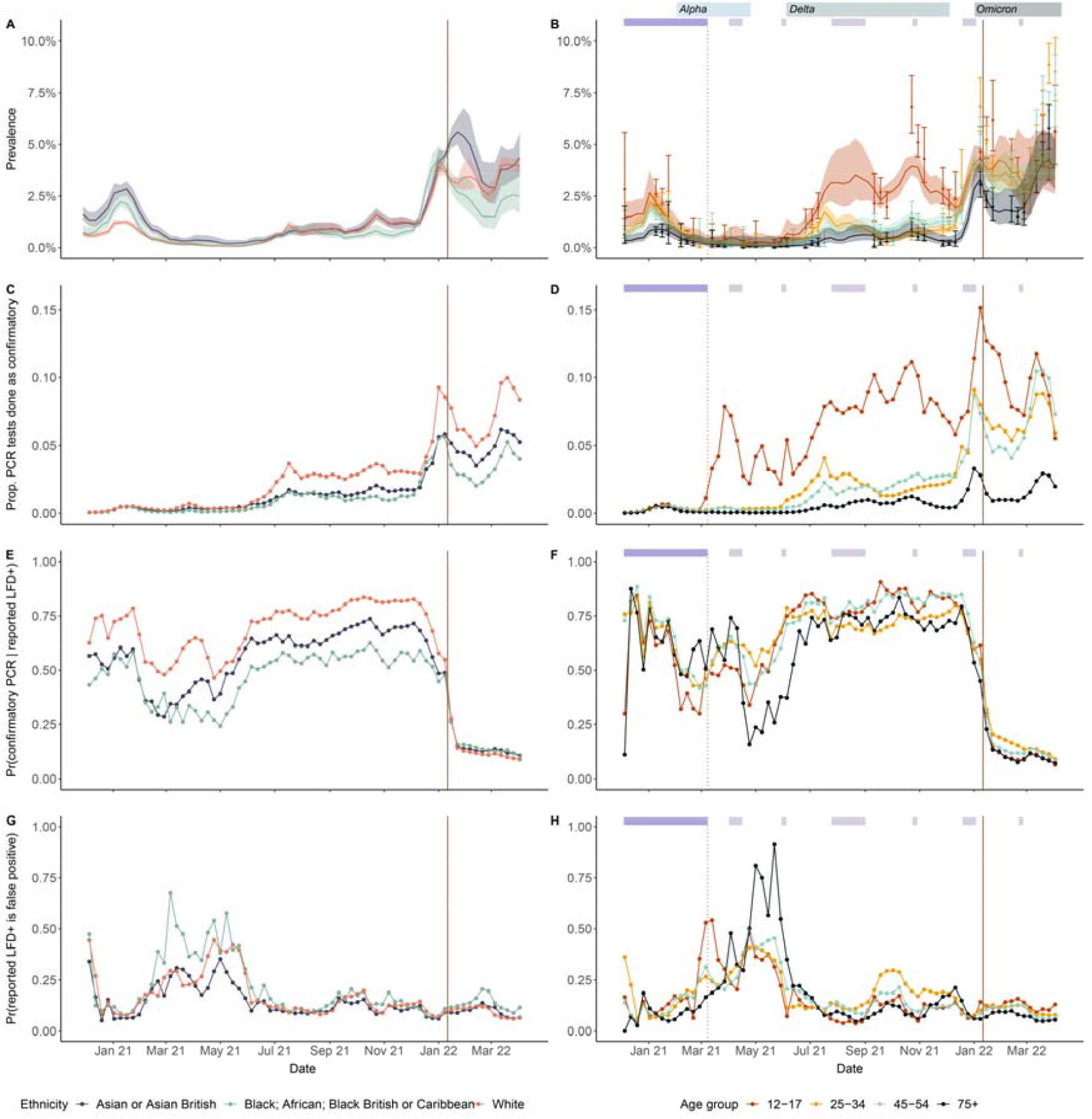
(A) Aggregated weekly national prevalence estimates for ethnic groups. This was the population-weighted average of posterior median estimates (solid lines) and 95% central credible intervals (shaded region) of ethnic group-specific debiased prevalences in LTLAs. (B) Aggregated weekly national prevalence estimates for age groups and 95% central credible intervals (solid and shaded areas respectively). National prevalence and 95% confidence intervals for age groups estimated by REACT-1 are shown as points and whiskers. (C, D) Proportion of PCR tests performed as confirmatory tests by ethnic and age groups. (E, F) Proportion of reported LFD positive tests followed by PCR confirmatory tests within 72 hours among different ethnic and age groups. (G, H) The probability of testing negative with a confirmatory PCR test after reporting a positive LFD test, among different ethnic and age groups, conditional on testing positive with an LFD, reporting the positive LFD test and seeking a confirmatory PCR test. The solid vertical line corresponds to the date (11^th^ January 2022) after which confirmatory PCR tests were no longer required following a positive LFD test, and the vertical dotted line corresponds to the date (8^th^ March 2021) when onsite asymptomatic testing was made available in schools. Restrictions on face-to-face education and school holidays are indicated by the dark and light purple rectangles, respectively. The blue rectangles represent the time periods of Alpha, Delta, and Omicron VOCs as defined in Methods. The date axis starts in December 2020, when LFD tests started being reported.

The roll-out of the universal testing offer sought to expand access to LFDs, but it corresponds with a separation in LFD testing patterns across ethnic groups (Figure 1H), and, subsequently, per capita LFD tests reported by White people were consistently higher than in other ethnic groups (from April 2021 per capita LFD tests were 34% (IQR: 22%, 44%) higher in White people versus Black people and 47% (IQR: 42%, 55%) higher in White people versus Asian people). Our estimates of prevalence in White people were similar to or lower than for other ethnic groups (Figure 2A) and similarly so for LFD positivity (Figure S8) suggesting that they performed and reported LFDs more frequently with asymptomatic or mild disease.

Working age populations had consistently the highest PCR testing coverages throughout the evaluation period (Figure 1I and Figure S9B) followed by that in the oldest group (those aged 75+). PCR testing coverages in secondary school-aged children (12-17) were more volatile, likely driven by school terms and the high variability in prevalence in these groups (Figure 2B). LFD test reporting trends were similar to those of PCR test seeking (Figure 1J), albeit with a large spike in LFD testing in school-aged children after the roll-out of asymptomatic testing in schools in March 2021^21^ (Figure 1A).

While the results described above are aggregated at the national level, there was variability between the sociodemographic groups at the LTLA level (Figure S11). This variation may be attributed to limited sample sizes, especially when we disaggregated the tests by self-reported ethnic groups in LTLAs.

From 11^th^ January 2022 onwards, confirmatory PCR tests were no longer required following a positive LFD test result due to the high rates of COVID-19 across the UK^16^, which explains the sharp decline in PCR tests in Figures 1E, G & I.

### Prevalence, false positivity and PCR positivity rate

During the Alpha wave, the SARS-CoV-2 prevalence in Asian people was considerably higher than for Black people which, in turn, was higher than for White people (Figure 2A), as was found previously^22^. During the Omicron BA.1 and BA.2 variant waves, the prevalence in Asian groups remained higher than for others, and this pattern was particularly striking in the North-West, West Midlands, and Yorkshire and the Humber regions (Figure S12). During the Omicron wave, the prevalence in White ethnic groups was higher than in Black ethnic groups. It is unclear what drove these complex prevalence patterns which remained generally consistent across regions of England, but natural and vaccine-induced immunity along with differences in mixing behaviour are possible explanations^23–25^.

SARS-CoV-2 prevalence in secondary school-aged children surged much higher than in other age groups during the VOC waves (Figure 2B), as has been reported previously^26,27^, likely driven by the higher contact rates of this age group. Prevalence in the oldest age group (75+) was generally lower than for others, although in early-mid 2022, it sharply increased and coalesced with the others, which may have been partly due to the perceived lower risk of severe disease from the Omicron variant, increased social mixing, and the generally high levels of vaccine coverage in this group^25,27^.

The proportion of PCR tests which were confirmatory varied substantially throughout the evaluation period and according to self-reported ethnicity and age group (Figure 2C&D). Generally, this proportion increased over time, and White people and secondary school-aged children typically had considerably higher proportions of PCRs which were confirmatory than other groups. In addition to asymptomatic testing in schools from March 2021 (Figure 1A), the lower risk of symptomatic disease^28^ and higher prevalence (Figure 2B) in this age group may partly explain their higher rates of confirmatory PCRs (Figure 2D).

Between October 2020 – December 2021, 60% reported LFD positives were followed up with confirmatory PCRs (Figure S6B). From December 2020 – December 2021, White populations were 1·45 times (IQR: 1·34, 1·51) more likely than Black populations to take PCR tests following a reported LFD positive and 1·33 times (IQR: 1·29, 1·47) more likely than Asian people (Figure 2E). Adherence to confirmatory testing policy was generally similar across age groups apart from those aged 75+ during the period between the end of the Alpha wave and the start of the Delta wave (Figure 2F). Figure 2G shows that from March – May 2021, the false positive rate (proportion of negative confirmatory PCRs) for Black populations was roughly 1·3 times (IQR: 1·07, 1·61) that in White people and roughly 1·85 times (IQR: 1·66, 2·00) that in Asian populations. From May – June 2021, the false positive rate was substantially elevated in those aged 75+ (Figure 2H). At least transiently then, there were times when there were substantial differences across ethnic groups in the risk of isolating unnecessarily if following the governmental policy to self-isolate. False positive rates are inversely related to disease prevalence, with the sensitivity and specificity of the test determining the exact relationship (see Supplementary Materials). This allowed us to estimate the sensitivity (range of central estimates: 66-72%) and specificity (99.5-99.9%) of LFDs by ethnicity and age group which we report in Table S5. While our posterior estimates had wide uncertainty intervals, the medians of these test characteristics were similar between age groups and differed across ethnic groups. These were in line with previous literature estimates^29,30^.

We also considered how the probability of a positive confirmatory PCR test declined with time elapsing between when the LFD positive was reported and the PCR test subsequently taken. We found that this rate of decay was considerably higher in Black people during the Alpha wave and generally remained above the other groups throughout the study period (Figure 3A), which could indicate that individuals in this ethnic group tended to test and report their positive LFD test later in their infection. Lower values of this parameter later in the study period may indicate that infections were caught earlier by LFD tests or may reflect a longer period of PCR detectability for later VOCs.

**Figure 3:**
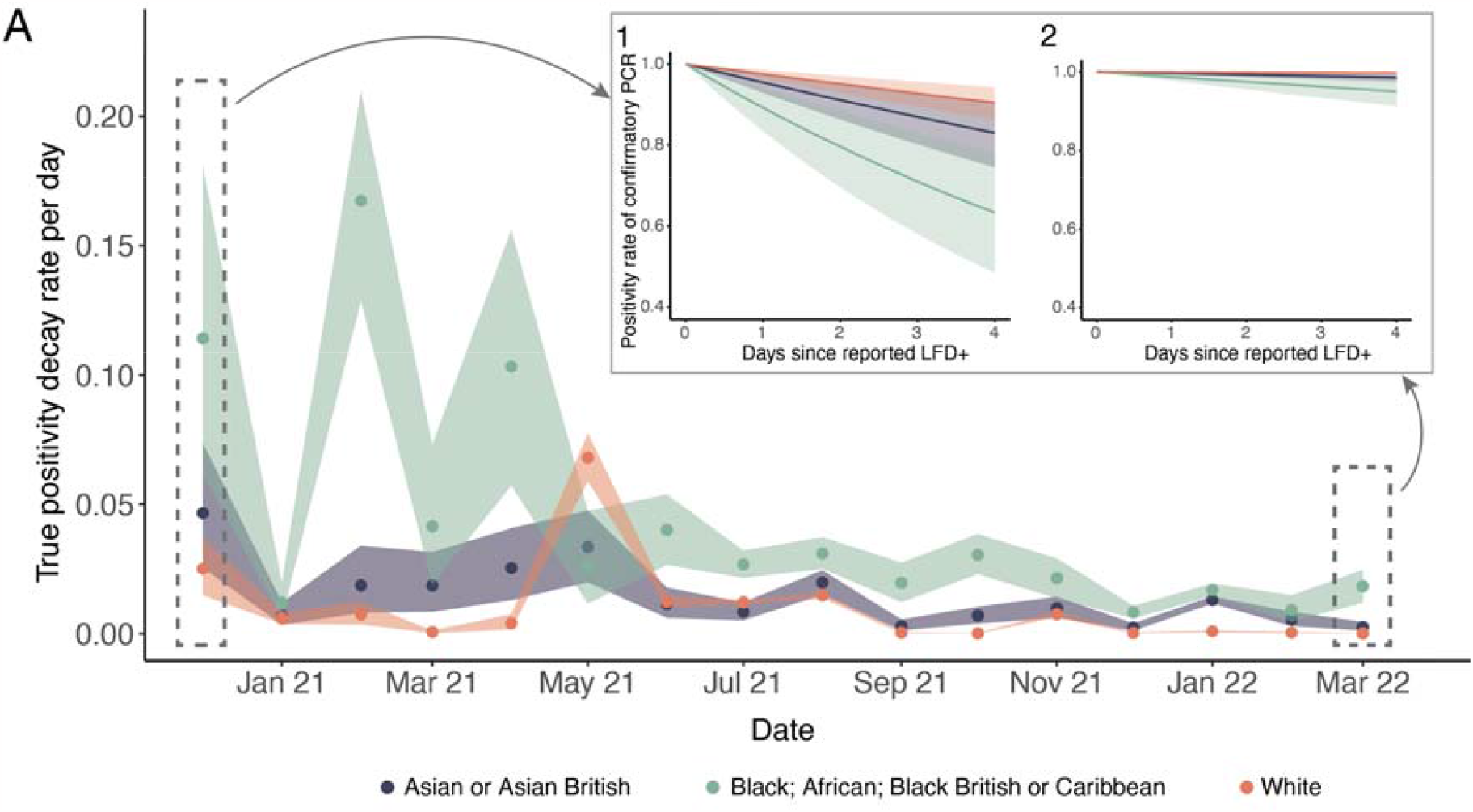
(A) Rate of decline in the probability of detecting a SARS-CoV-2 infection by PCR within 3 days from reporting a positive LFD test over the study period by ethnic groups. Solid dots (and shaded areas) in the main figures are the posterior median estimates (and 50% central credible intervals) of ethnic group specific decay rates. The inset plots translates what the rate estimates in A correspond to in terms of the rate of decline in probability of testing positive by PCR subsequent to reporting of an LFD positive (with 50% credible intervals) for the first (1: December 2020) and last month in our study (2: March 2022).

### Trends in testing biases

Our analysis indicates that infected individuals were much more likely to be tested by PCR than those uninfected. The testing biases for LFDs were considerably lower than for PCRs, meaning symptoms played a lesser role in shaping testing behaviours for LFDs. These results for both PCR and LFD tests are reflective of national testing policy throughout the study period. Throughout the analysis period, the relationship between testing biases (Figure 4C&D) and prevalence (Figure 2A&B) was not strong, but during the Delta wave and Omicron waves, there were visible increases in PCR testing biases (Figure 4C). In February 2022, the government removed the legal requirement for those in England to self-isolate following a positive test result and for close contacts who were fully vaccinated to conduct daily tests^31^. It is possible that this policy change prompted a shift towards increased usage of LFDs for symptomatic testing, which could explain the increasing bias in the use of LFDs in infected individuals (Figure 4C, D).

**Figure 4:**
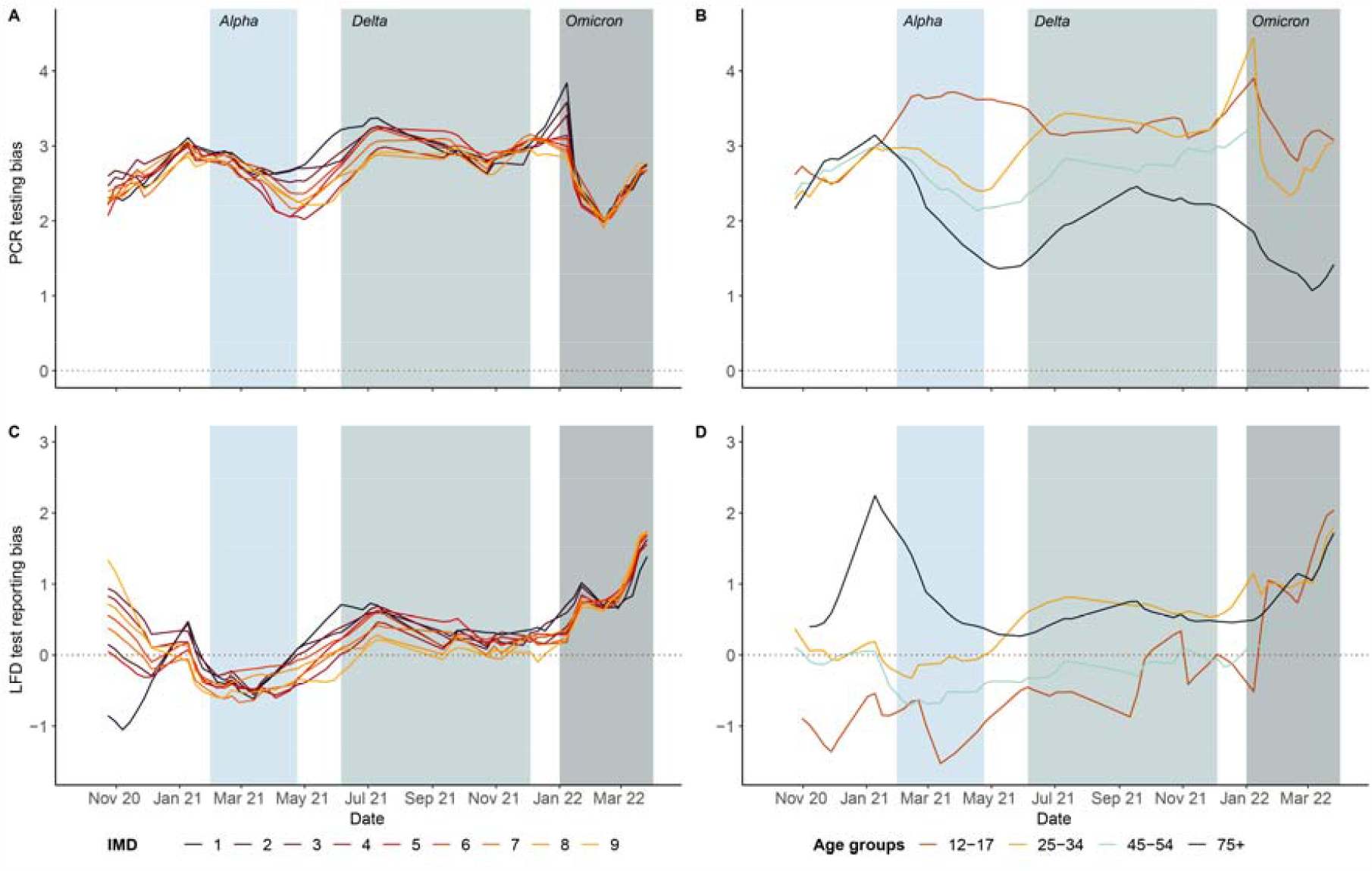
(A, B) Estimated mean PCR testing bias (log odds of taking a PCR test in infected versus uninfected subpopulations) by deprivation levels and age groups in England over time. (C, D) Estimated mean LFD test reporting bias in deprivation levels and age groups. 1 and 9 corresponding to most deprived and least deprived levels respectively. Shaded areas from left to right, correspond to the time periods of Alpha, Delta, and Omicron VOCs respectively (defined in Methods). Testing biases with uncertainty intervals and other age groups are provided in Figure S13, S14.

We found that deprivation and age were associated with testing biases on average, however the uncertainty intervals overlapped across deprivation levels (Figure S13), although the age specific patterns were more distinct (Figure S14). During the invasion phases of the Delta and Omicron VOCs, the most deprived populations were disproportionately more likely to test themselves via PCR only when infected versus when uninfected, and the same pattern held for LFD test-then-reporting. For example, in June 2021, at the start of the Delta wave (the second shaded box in Figure 4A), the average odds of PCR testing in the most-deprived areas were exp(3·21) ≈ 24 times higher in individuals with infection compared with individuals without infection; at the same time, the corresponding figure was only ≈ 11 in the least-deprived areas. A range of hypotheses could explain these behavioural differences, such as differential costs of self-isolation, differences in test accessibility and different views about the risks posed by infection, but our data cannot probe these. From mid-February 2022, individuals in areas with different levels of deprivation had more similar PCR test-seeking behaviour. This was likely due in part to the lifting of the legal requirement to self-isolate^31^.

Starting with the Alpha wave, the infection status for those aged 75+ played a lesser role in determining whether individuals tested themselves via PCR than for younger age groups (Figure 4B). This could be because older individuals were more at risk of severe disease, and they tested themselves more frequently by PCR due to caution^17^. It could also be because older people generally have more comorbidities, some of which may produce symptoms similar to COVID-19 causing them to seek PCR tests. We also found that the PCR testing bias was particularly high in school aged children (12-17 years) groups during March –June 2021 (Figure 4B) when LFD was extensively being used (corresponding to a low bias in LFD in Figure 4D)^21^. LFD test reporting biases were more similar across age groups, although the oldest age group had a higher bias early on during the analysis period (Figure 4D).

### Incidence and case detection ratio trends

By comparing the estimated true underlying incidence (using REACT-1 and ONS estimates) to reported cases (defined as PCR and LFD positive tests), we found that the national testing programme detected between 26%-40% of all cases across the entire evaluation period (see Methods), although this was likely an overestimate because of potential double counting of LFD and confirmatory PCR tests in the Pillar 2 data as well as multiple tests by the same person. This ratio varied substantially over time and increased until January 2022 after which PCR tests were no longer mandatory following a positive LFD test^16^. While the differences in the REACT-1 and ONS derived ratios (Figure 5A) are likely explained by the differences in their sampling strategies (and prevalence estimates), there were no notable persistent differences in case detection ratios according to the deprivation levels of LTLAs (Figure 5B). Throughout much of 2021, Black ethnic groups had higher rates of case detection (Figure 5C), and it is unclear what underpins this relationship, but targeted community testing could have played a role^19,20^.

**Figure 5:**
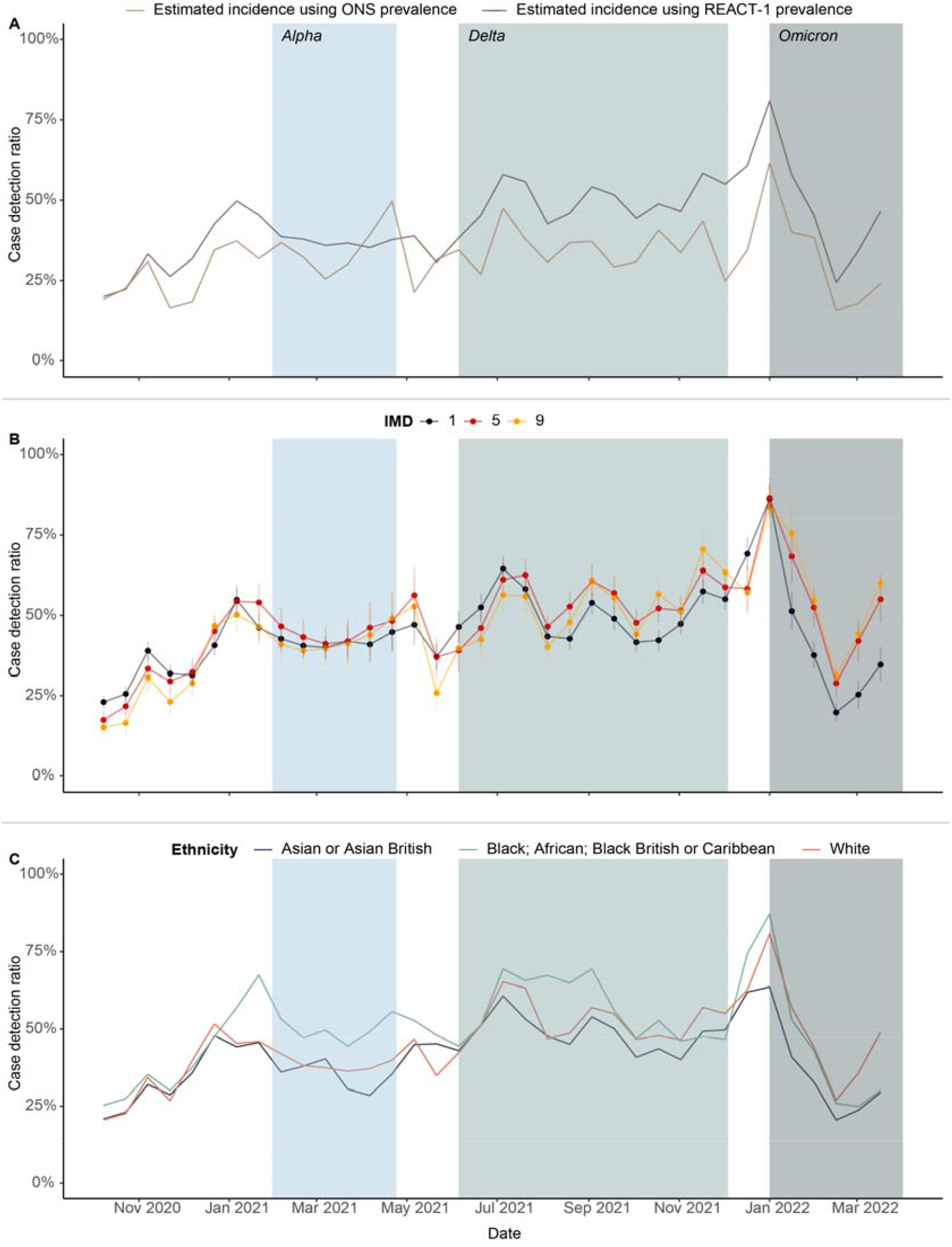
(A) Bi-monthly national estimated case detection ratio over time, using PCR and LFD positive and estimated incidence. The dark and light brown curves represent ratios using REACT-1 and ONS prevalence data, respectively. (B) Case detection ratio over time by deprivation levels (IMD). The dots (and error bars) represent the coefficient estimates (and standard error) of a linear regression with LTLA specific case detection ratio as the dependent variable and IMD levels as predictors for each month. Here IMD level 1 (black) and 9 (yellow) refer to the most- and least-deprived areas, respectively. (C) Bi-monthly national case detection ratio tests by ethnic groups. Shaded areas from left to right, correspond to the time periods of Alpha, Delta, and Omicron VOCs respectively (defined in Methods).

## Discussion

Our work reveals the intricate and dynamic nature of COVID-19 testing patterns in England throughout our study period (October 2020 - March 2022), illustrating strident differences in test use and reporting between sociodemographic groups. These differences complicate and obfuscate the real-time assessment of transmission across non-traditionally reported groups, and our prevalence estimates point towards an unequal burden of infection across them.

Importantly, these insights were based on the PCR testing programme available to the population and the universal testing offer for LFD tests. However, the LFD tests were also accessible to individuals through a variety of other routes such as school, university, workplace or adult social care settings and we do not distinguish between these. In addition to these two, the targeted community testing service, established in July 2021, aimed to reach disproportionately impacted people and those underserved by the national universal testing offer^19,20^. These target populations included those in areas of social economic deprivation; ethnic minority groups; those in high-risk occupations; individuals experiencing homelessness or who were sleeping rough; migrants; asylum seekers; refugees; and Gypsy, Roma and Traveller communities. This service was designed and delivered by local authorities and their partners which aimed to more directly reach disproportionately impacted groups. Unfortunately, while tests from this service were included in the data we analysed, targeted testing data were not separately labelled, and we could therefore not summarise this service’s data in isolation.

We considered three categories of sociodemographics in this work: self-reported ethnicities of individuals testing or reporting test results; their self-reported age; and a grouping of individuals according to the index of multiple deprivation of the LTLA where each test was taken (PCR) or taken and reported (LFD). We chose these categories of analysis opportunistically according to criteria recorded in the testing data. These are suboptimal because they pool people according to somewhat arbitrary characteristics meaning they lack operational use; if it is known that self-reported Black individuals have a higher burden of infection, how should public health authorities actually respond? This group encompasses a huge diversity of different groups who live and interact in different communities. Existing categorisations also miss undocumented and presumably vulnerable individuals, such as undocumented migrants. Furthermore, there is compelling evidence suggesting a connection between belonging to an ethnic minority group and residing in economically disadvantaged areas, leading to an unequal burden of COVID-19^32^. While our current study examines these factors in isolation, a potential future expansion could explore their combined impact^22^. Social scientists and local community leaders are better placed to understand these nuances, and interdisciplinary collaborations between these and those working in public health should be prioritised^33,34^.

The CORSAIR study, conducted between March 2020 and January 2021, found that financial hardship, IMD, lower socioeconomic status and having a dependent child in the household were associated with poorer recognition of symptoms and a lower likelihood of booking a test^35^. Major barriers to testing for people in deprived areas were the financial and practical implications of having to self-isolate following a positive COVID-19 test^18^, and those from ethnic minority backgrounds were disproportionately affected by this^36^. These barriers were also acute for those working in sectors which effectively closed during the pandemic, and women and young people were more likely to fall into this group^37,38^. The Test and Trace Support Payment scheme^39^ aimed to provide a financial safety net for those required to self-isolate, but many in need had difficulties accessing the scheme^40^.

The PCR and LFD data from Pillar 2 which we explore offer complementary viewpoints of testing uptake and behaviour since PCR test results reflect test uptake (as the taking of a test resulted in the data being logged failing any mistakes in data recording) whereas LFD test data were available only if individuals self-reported their results. We find that the use (and for LFD, reporting of) these two tests were different throughout the study period, with PCR test use heavily concentrating in those infected and reported LFD positivity giving a more representative view of underlying prevalence; both of these are supportive of the general testing policy in England throughout the period. However, only 15% of LFD tests distributed were reported^9^, and, unfortunately, data on subnational test distribution were unavailable, which prevented granular analysis of LFD uptake and reporting.

During infectious disease epidemics, testing can guide individual- and community-level interventions, for example by allowing self-isolation following exposure, and a return to work following a negative test result. Community surveillance surveys such as the world-leading REACT-1 and ONS-CIS surveys rolled-out in England and the UK, respectively, used representative samples of individuals to produce estimates of SARS-CoV-2 infection prevalence, while adjusting for selection bias due to non-response^41,42^. Ours and previous work^15^ has shown how these high-quality, low-volume surveys can be combined with mass testing data to determine fine scale estimates of disease prevalence for various spatial scales or sociodemographic groups. These data could then be used to determine granular estimates of disease transmission, such as the time-varying reproduction number, R_t_. Such nowcasting approaches could be deployed in real-time in future pandemics to improve disease surveillance, particularly to probe transmission in minority risk groups and spatial areas for which community surveillance random sampling surveys in isolation are insufficiently powered to estimate accurately.

### The EY-Oxford Health Analytics Consortium membership list

Ricardo Aguas, Ma’ayan Amswych, Billie Andersen-Waine, Sumali Bajaj, Kweku Bimpong, Adam Bodley, Liberty Cantrell, Siyu Chen, Richard Creswell, Prabin Dahal, Sophie Dickinson, Sabine Dittrich, Tracy Evans, Angus Ferguson-Lewis, Caroline Franco, Bo Gao, Rachel Hounsell, Muhammad Kasim, Claire Keene, Ben Lambert, Umar Mahmood, Melinda Mills, Ainura Moldokmatova, Sassy Molyneux, Reshania Naidoo, Randolph Ngwafor Anye, Jared Norman, Wirichada Pan-Ngum, Sarah Pinto-Duschinsky, Sunil Pokharel, Anastasiia Polner, Katarzyna Przybylska, Emily Rowe, Sompob Saralamba, Rima Shretta, Sheetal Silal, Joseph Tsui, Merryn Voysey, Marta Wanat, Lisa White, Gulsen Yenidogan

## Data Availability

The data were made available to the paper authors by UKHSA as part of a retrospective evaluation of England's COVID-19 testing programme. The authors cannot make the underlying datasets publicly available for ethical and legal reasons particularly due to the sensitive information included. Applications for relevant anonymised data should be submitted to UKHSA. The code for the statistical models is available from our Github repository.

https://github.com/sumalibajaj/equitytesting

## Acknowledgements

The authors would like to express gratitude to Oliver Munn, Sarah Tunkel, Nick Sharp, Sariyu Shoge and Olutoye Olatunbosun of UKHSA for sponsoring this research and enabling access to the data used in this study. We thank the REACT investigators for their assistance and for access to data from the REACT-1 study. REACT-1 was funded by the Department of Health and Social Care. This study was funded by Secretary of State for Health and Social Care acting as part of the Crown through the UK Health Security Agency (UKHSA), reference number C80260/PRO5331. S.B. is supported by the Clarendon Scholarship and St. Edmund Hall College, University of Oxford and NERC DTP [grant number NE/S007474/1]. C.A.D. is supported by the UK National Institute for Health Research Health Protection Research Unit (NIHR HPRU) in Emerging and Zoonotic Infections in partnership with Public Health England (PHE), Grant Number: HPRU200907.

