## Supplementary material for "Understanding COVID-19 testing behaviour in England through a sociodemographic lens"

Supplementary Information

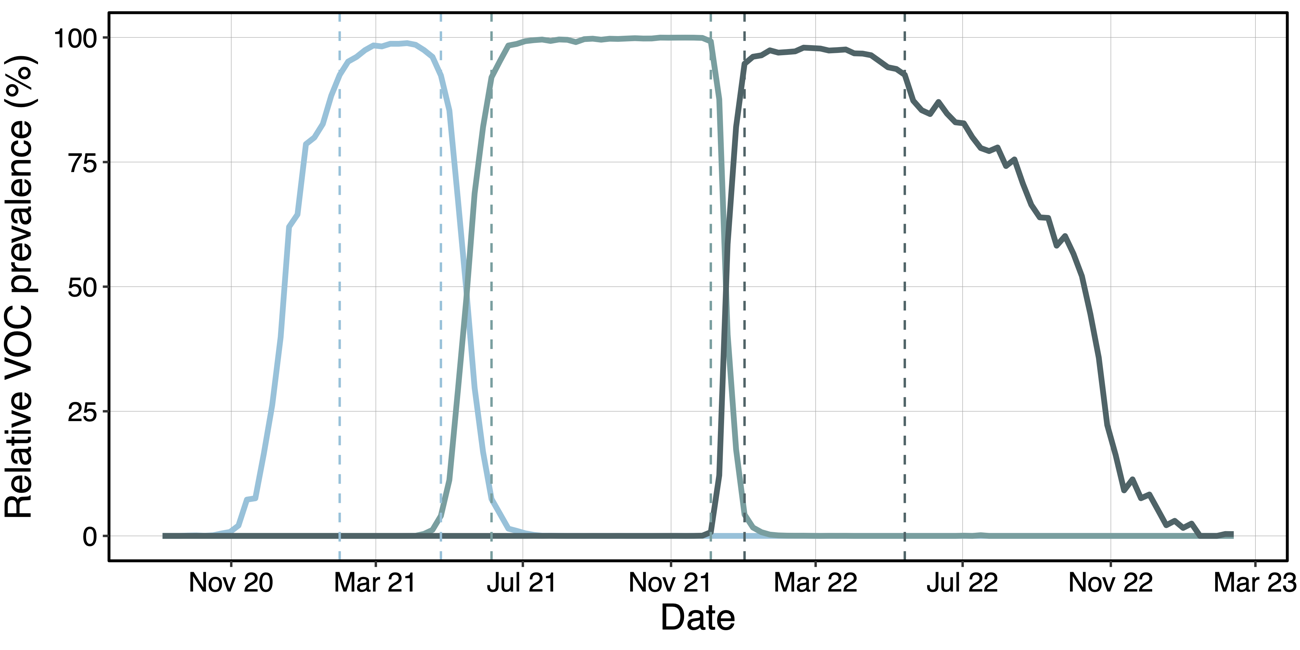

**Figure S1: Variant time periods.** Time periods during which Alpha, Delta and Omicron (left-to-right in figure) variants of concern (VOC) were dominant (>= 90% relative prevalence) in England. The solid curves are the VOC specific weekly proportions using lineage counts from Wellcome Sanger Institute^1^ and the dashed coloured lines delineate the time periods when the relative proportion of each of the VOCs equalled or exceeded 90%.

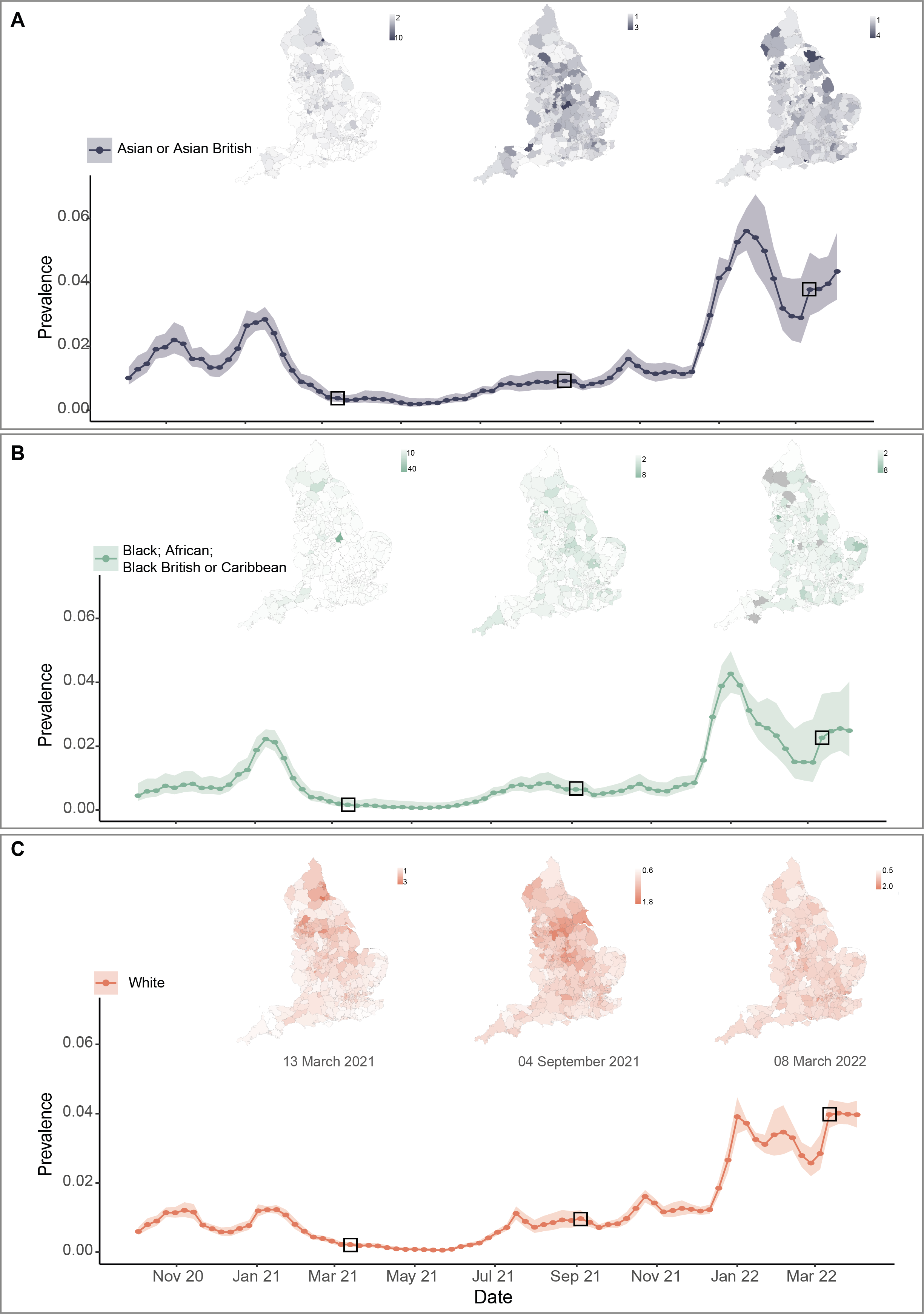

**Figure S2: Prevalence of SARS-CoV-2 in ethnic groups.** (A) Asian ethnic group (B) Black ethnic group, and (C) White ethnic group over time calculated using a weighted mean of our fine scale prevalences and 95% central credible intervals of ethnic groups within LTLAs and their respective population sizes. The maps show the age group specific prevalences between LTLAs during the mid-periods of Alpha, Delta, and Omicron VOCs as defined in Supplementary methods section “Variant time periods”. The map shading is given by the ratio between the prevalence of an ethnic group in an LTLA and the national ethnic group specific prevalence during that week, indicating deviations from the average country-wide prevalence.

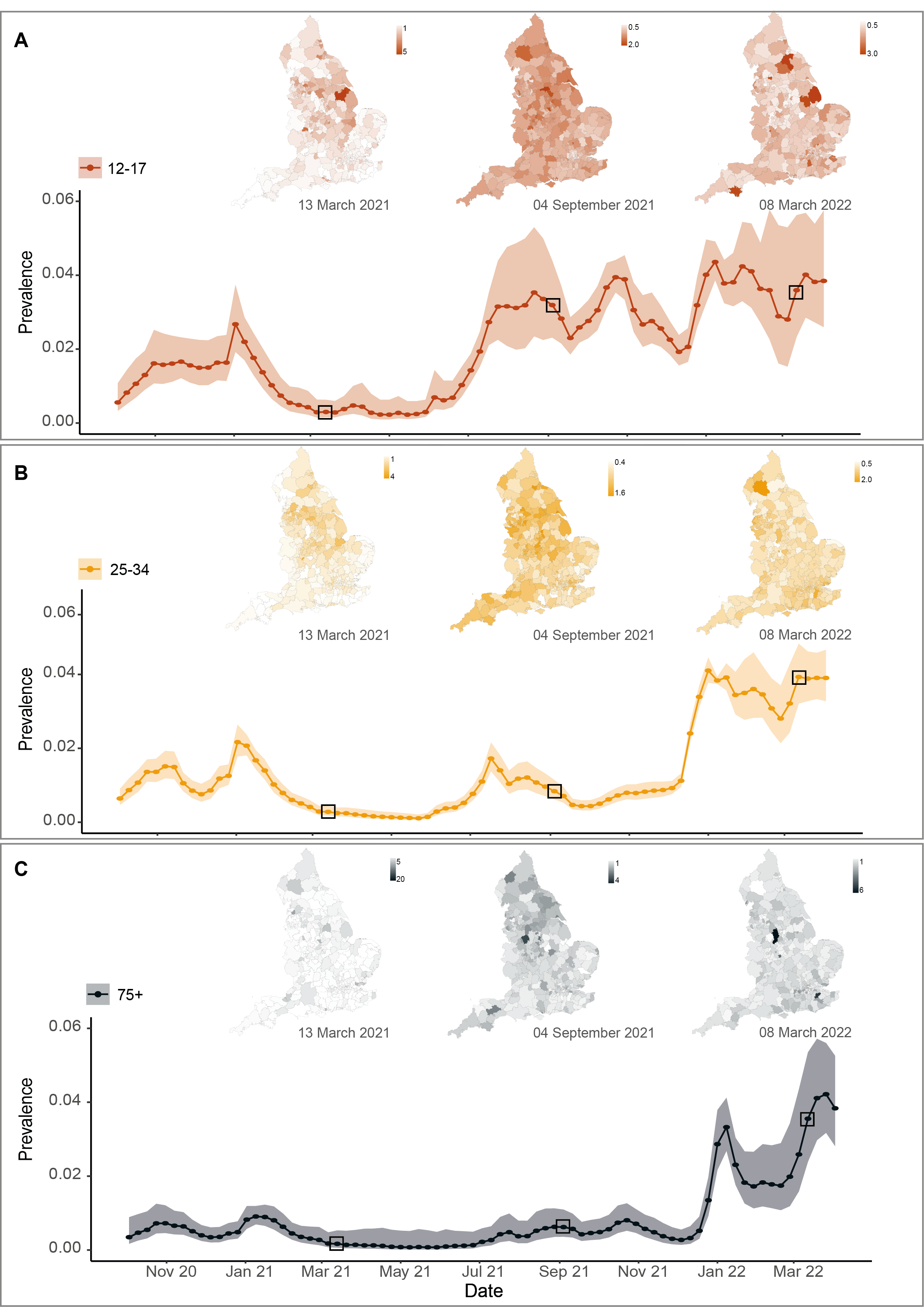

**Figure S3: Prevalence of SARS-CoV-2 in individuals by (self-reported) age groups. (**A) 12-17 years (B) 25-34 years, and (C) 75+ years over time calculated using a weighted mean of our fine scale prevalences and 95% central credible intervals of age groups within LTLAs and their respective population sizes. The maps show the age group specific prevalences between LTLAs during the mid-periods of Alpha, Delta, and Omicron VOCs as defined in Supplementary methods section “Variant time periods”. The map shading is given by the ratio between the prevalence of an age-group in an LTLA and the national age-group specific prevalence during that week, indicating deviations from the average country-wide prevalence.

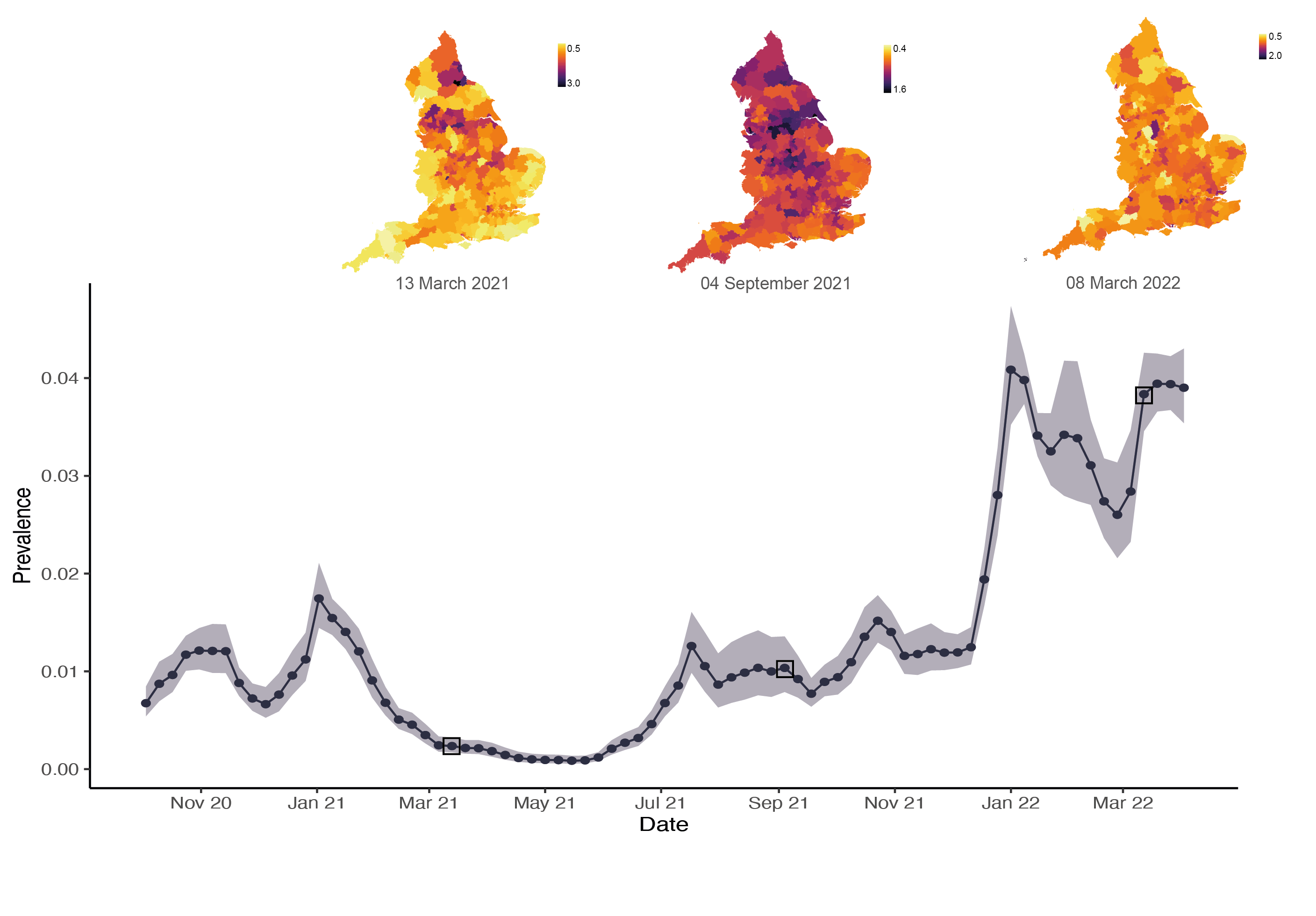

**Figure S4: Prevalence of SARS-CoV-2 in England over time and by LTLA.** Using a weighted mean of our fine scale LTLA prevalences and 95% central credible intervals and their respective population sizes. The maps show the LTLA-specific prevalences during the mid-points of Alpha, Delta, and Omicron VOCs as defined in Supplementary methods section “Variant time periods”. The legend for the maps indicates a scale for the ratio between the prevalence of an LTLA and the national prevalence during that week, indicating which areas were higher or lower than the average country-wide prevalence.

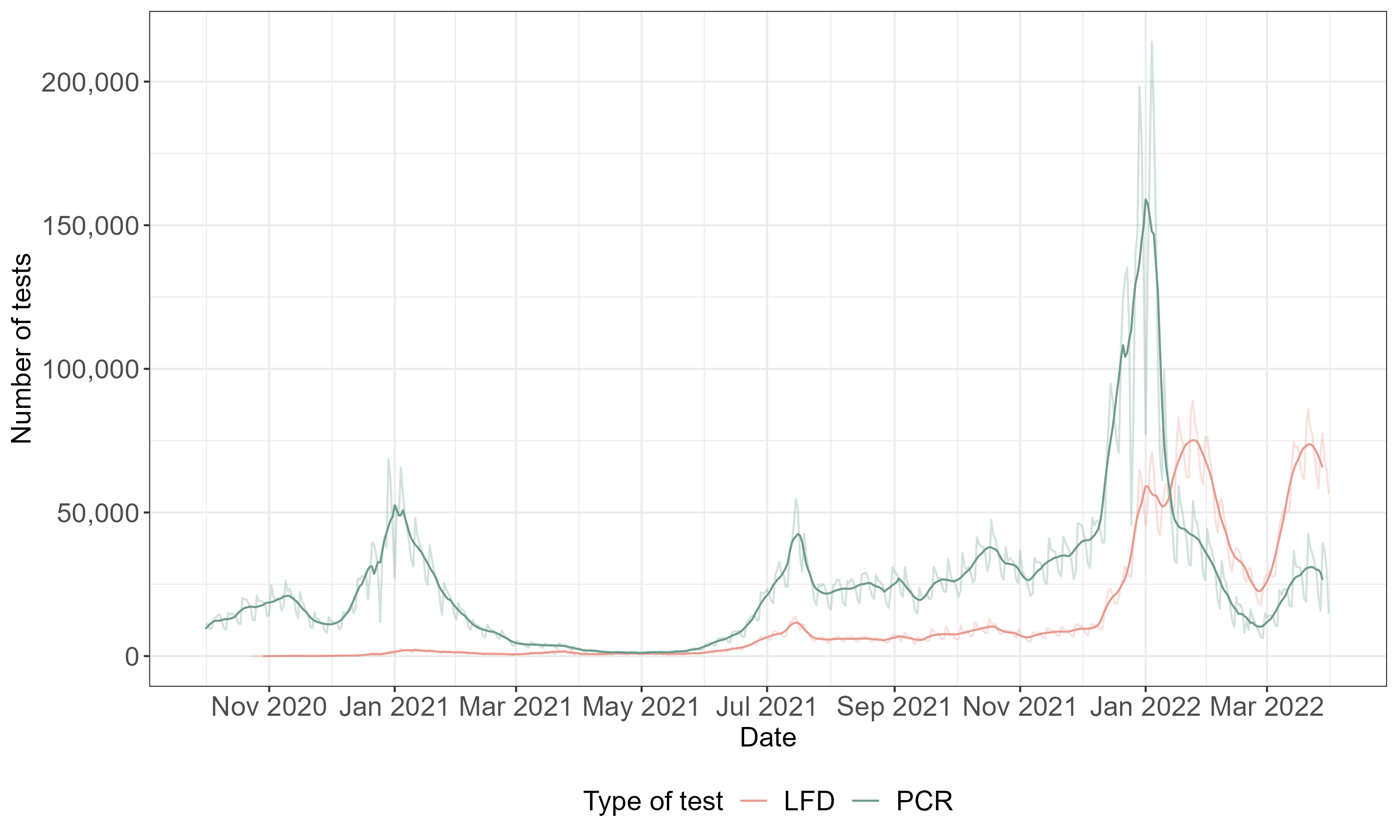

**Figure S5: Number of PCR tests performed and LFD tests reported in England.** The light-coloured curves are the raw numbers, and the darker curves display data that has been smoothed using a 7-day moving average (centred on the current day).

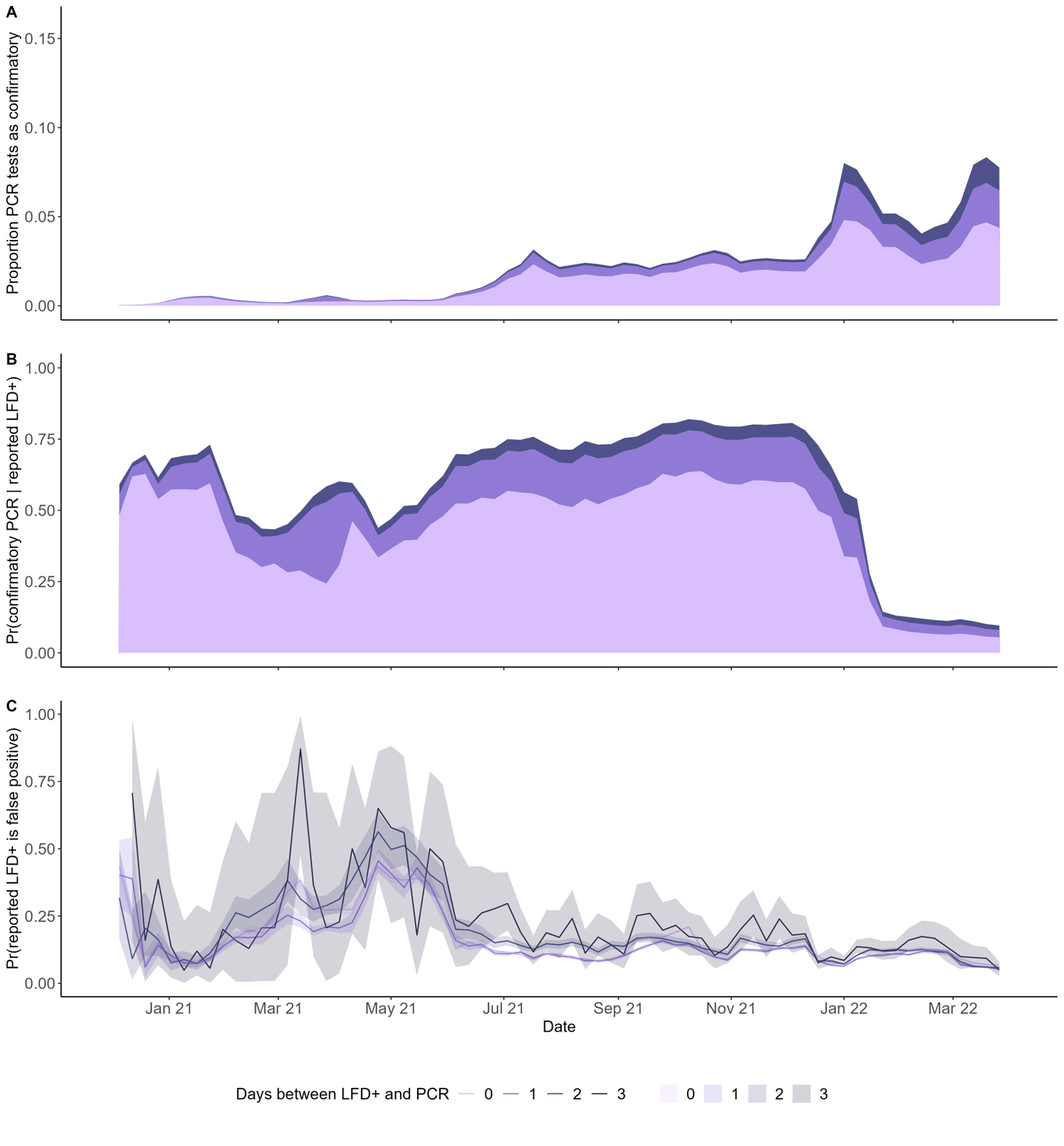

**Figure S6: Confirmatory PCR tests over time.** (A) Proportion of PCR tests performed as confirmatory tests within given number of days since reporting an LFD positive result. (B) Proportion of reported LFD positive tests followed by PCR confirmatory tests within given number of days. Both A and B are stacked charts, which show how the proportions and probabilities, respectively, increase as the window between the LFD test being reported and the PCR test occurring increases. (C) Posterior median probability of testing negative with a confirmatory PCR test after reporting a positive LFD across the study period and dependent on the time from when the LFD positive test was reported and the PCR test taken. The uncertainty intervals represent 95% central credible intervals, calculated assuming a uniform prior on the probability of a false positive.

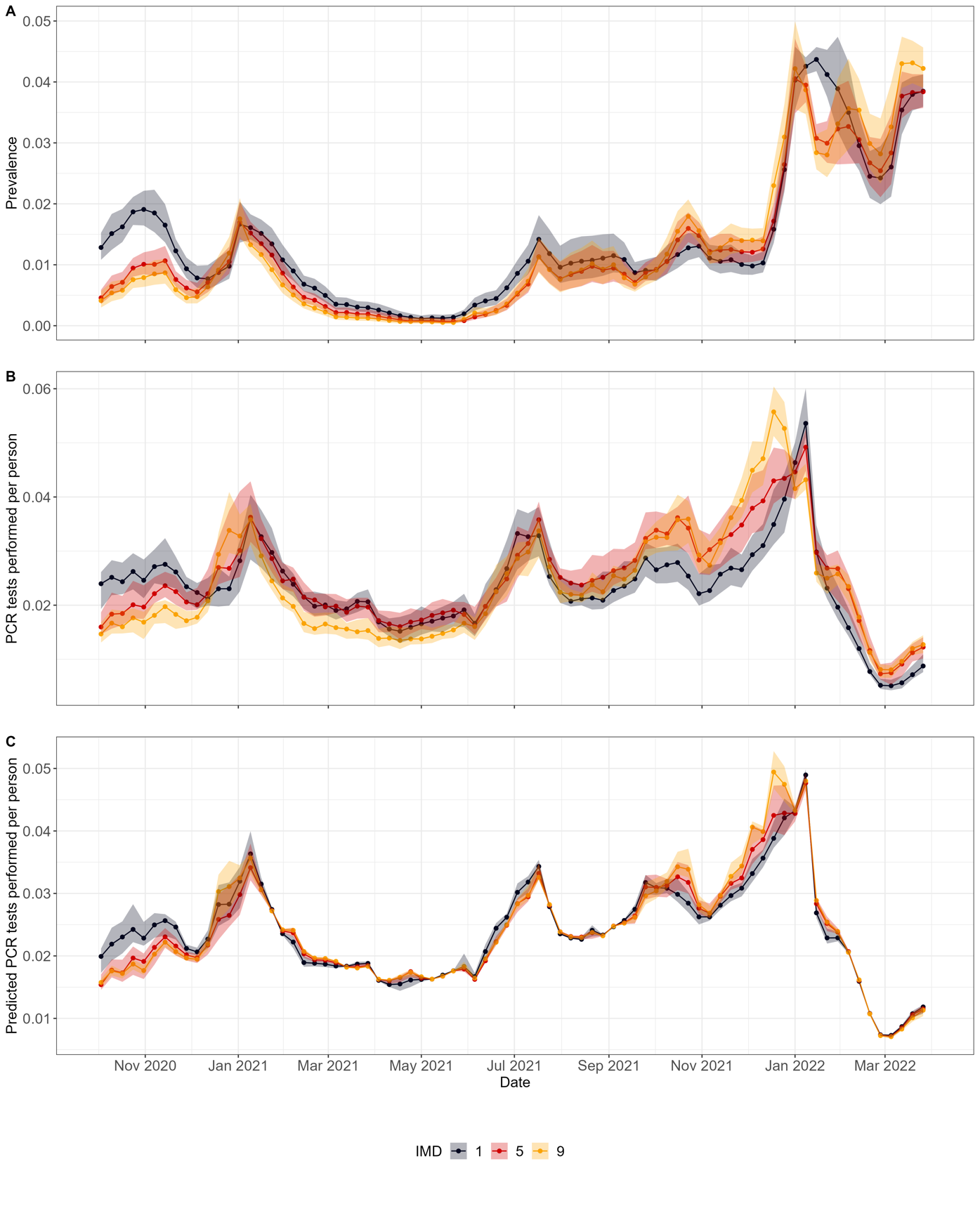

**Figure S7: Relationship between deprivation, SARS-CoV-2 prevalence and PCR tests performed.** (A) Aggregated weekly national prevalence estimates for IMD levels (1=most deprived, 9 = least deprived). This was calculated by weighing the LTLA-specific debiased prevalences and 95% central credible intervals by their population sizes into IMD levels. The solid curves are the posterior medians and the shaded areas are the 95% central credible intervals. (B) Observed weekly median volume of PCR tests per capita per LTLA by three levels of IMD and shaded regions indicate the interquartile range across the LTLAs. (C) Predicted PCR volumes from weekly regressions between LTLA specific prevalences and PCR volume. See Supplementary methods sub-section “LTLA prevalence and PCR tests” within the section “Causal debiasing approach for obtaining time-varying estimates of SARS-CoV-2 prevalence at fine scale resolution and for estimating testing biases” for details about the analysis. The solid curves show the median volume of predicted PCR tests and shaded areas indicate the interquartile range across all LTLAs according to their IMD level.

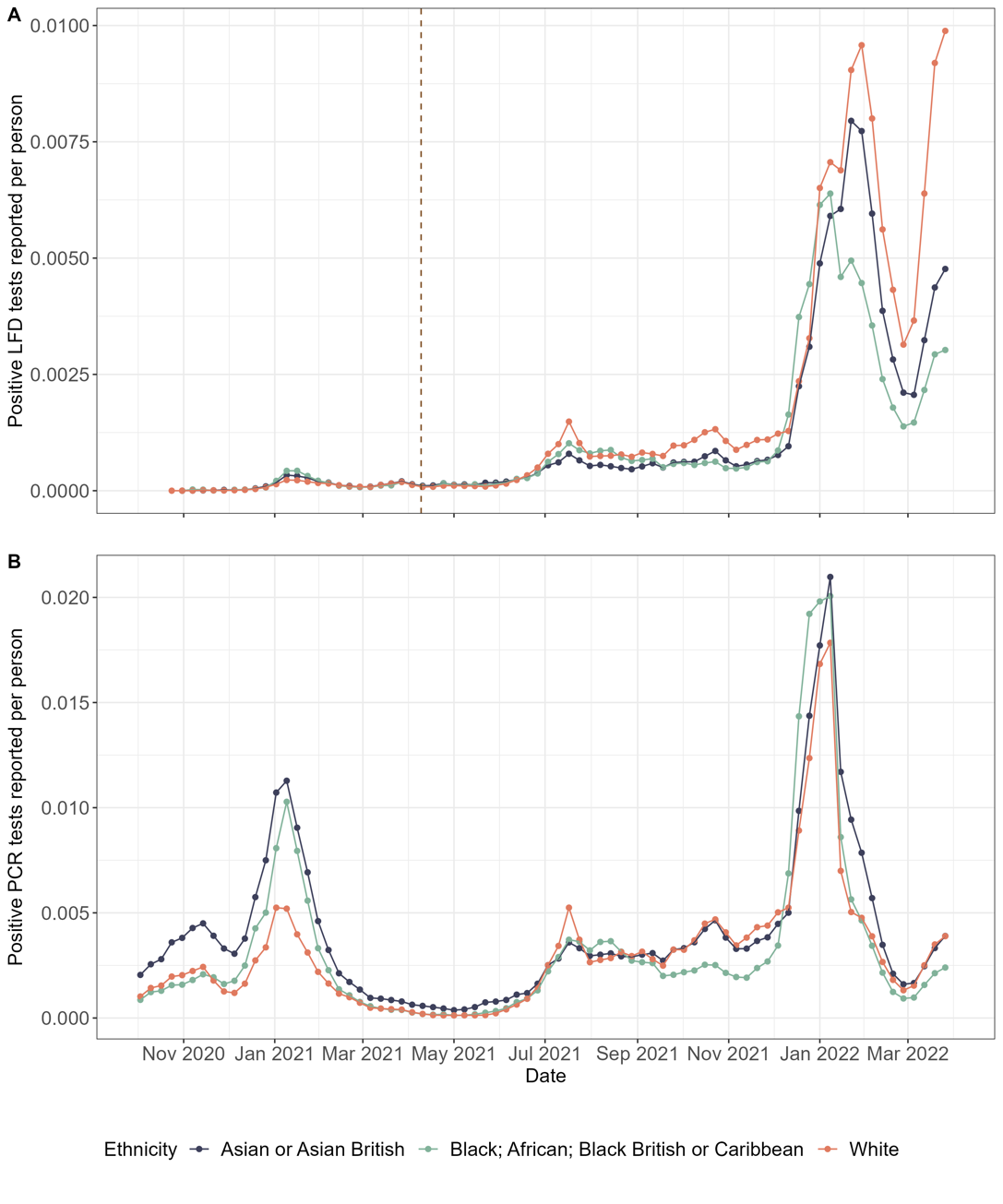

**Figure S8: PCR and LFD positive test performed/reported per capita by ethnic groups.** The vertical dashed line corresponds to the date (9^th^ April 2021) when free asymptomatic testing was made available to all of England.

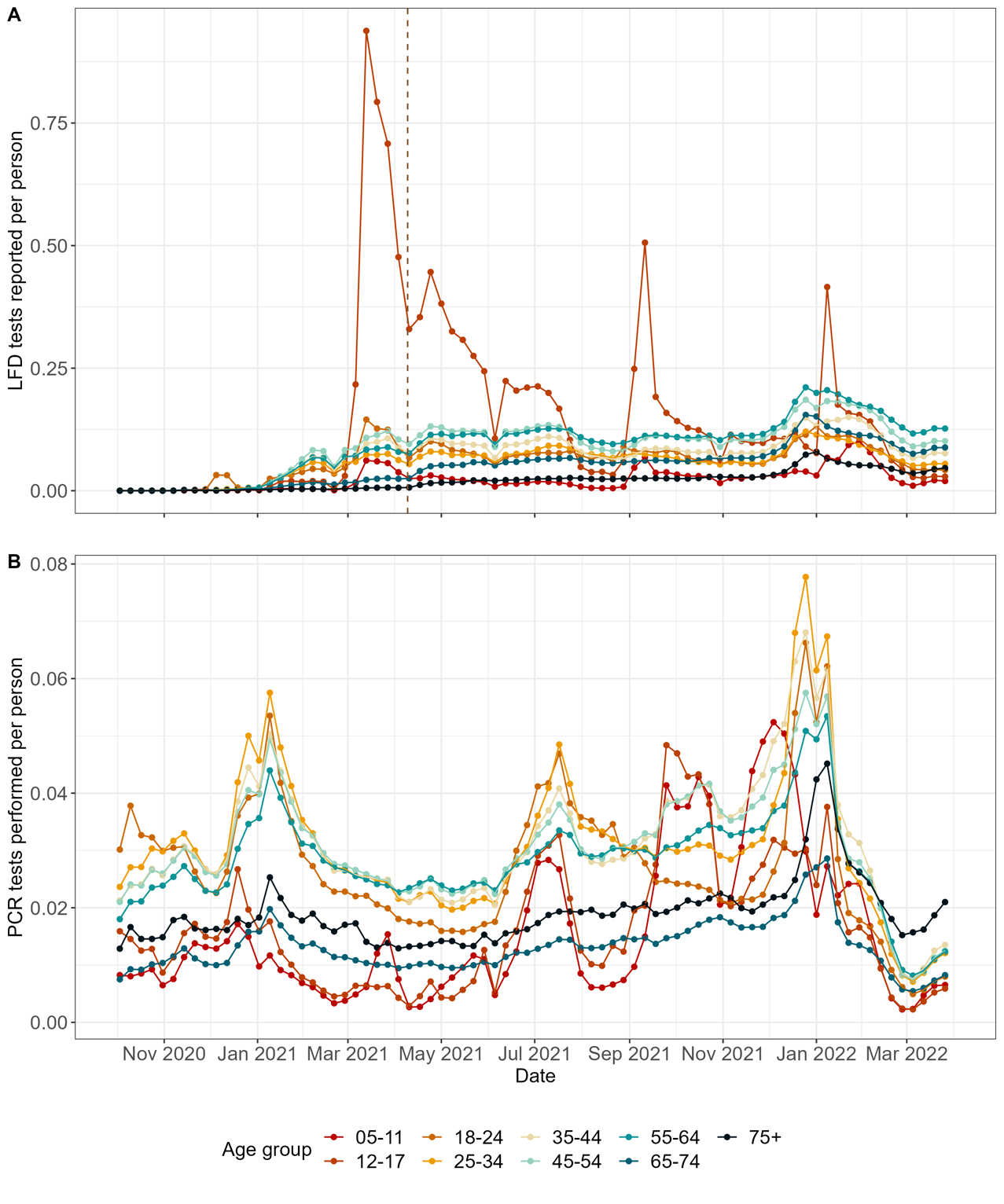

**Figure S9: Weekly number of PCR and LFD tests performed/reported per capita by age groups in England.** The vertical dashed line corresponds to the date (9^th^ April 2021) when free asymptomatic testing was made available to all of England.

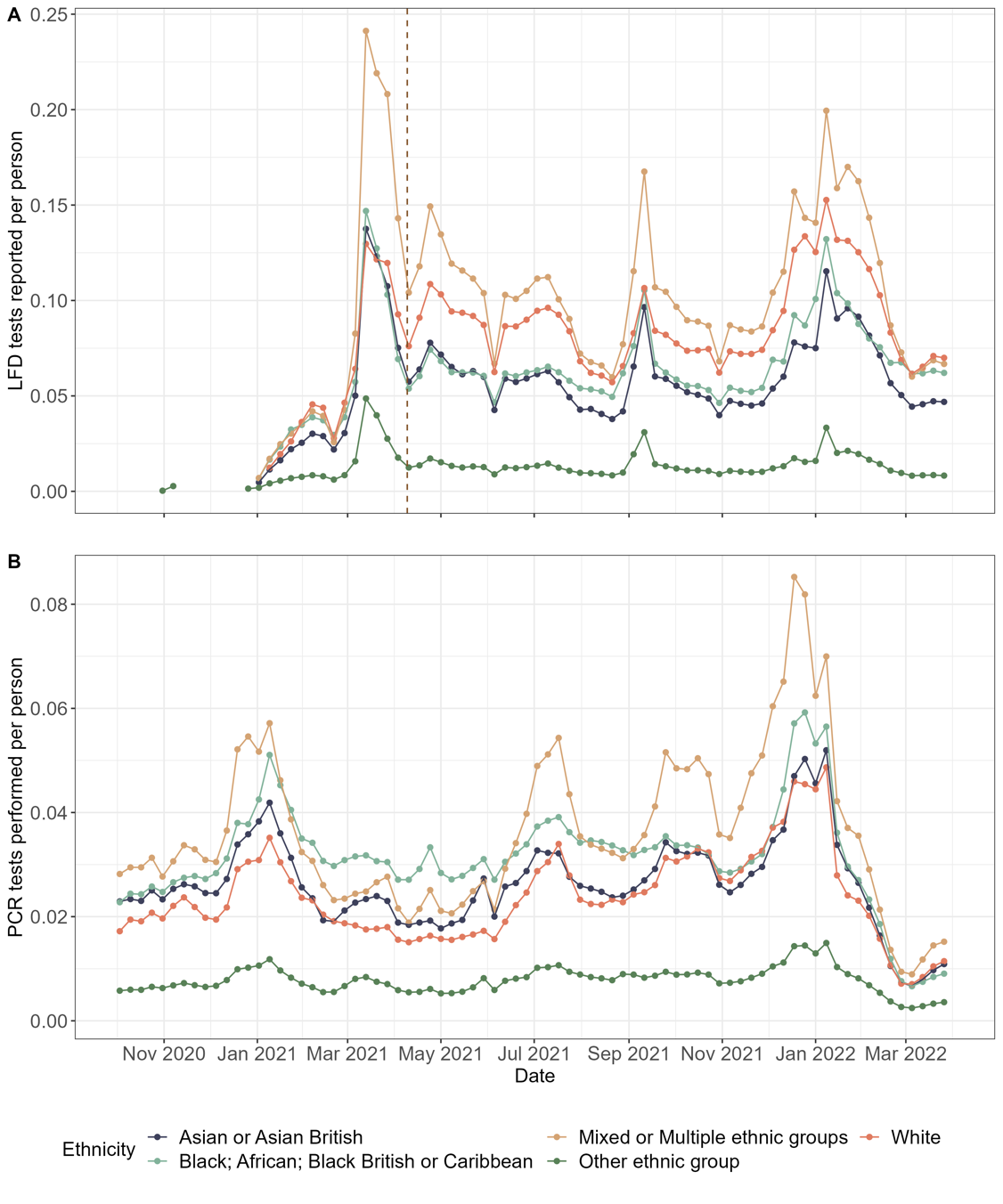

**Figure S10: Weekly number of PCR and LFD tests performed/reported per capita by five ethnic groups in England.** The vertical dashed line corresponds to the date (9^th^ April 2021) when free asymptomatic testing was made available to all of England.

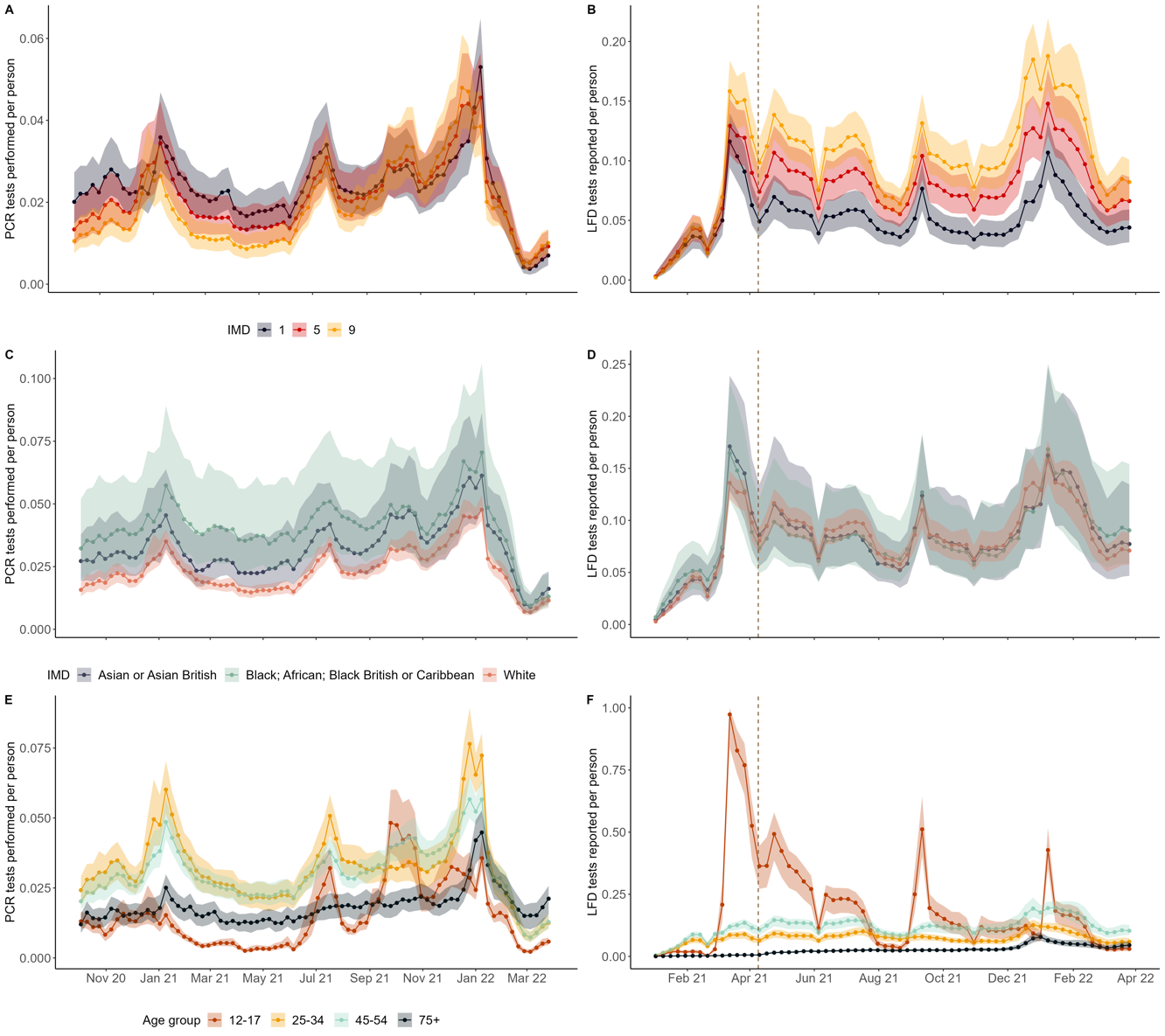

**Figure S11: Aggregated LTLA- and sociodemographic group-specific PCR tests performed and LFD tests reported per person.** For each LTLA the number of PCR tests performed per person and LFD tests reported per person are calculated by (A, B) deprivation level, (C, D) ethnicity, and (E, F) age group. The solid lines and dots represent the median tests per person by each subgroup and shaded regions indicate the interquartile range across all LTLAs.

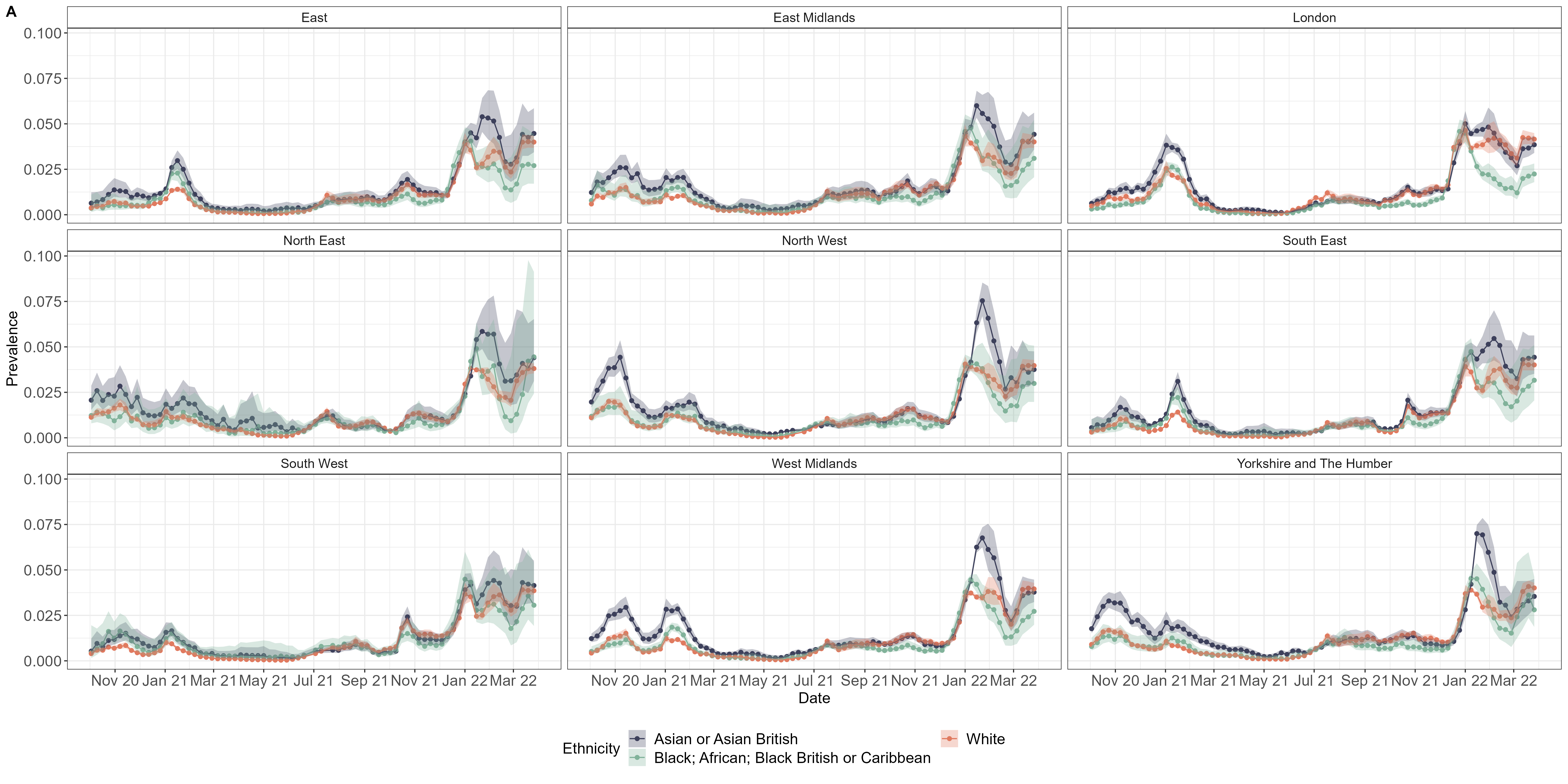

**Figure S12: Weekly regional prevalence estimates by ethnic group.** These were population-weighted average of posterior median estimates (solid curves) and 95% central credible intervals (shaded regions).

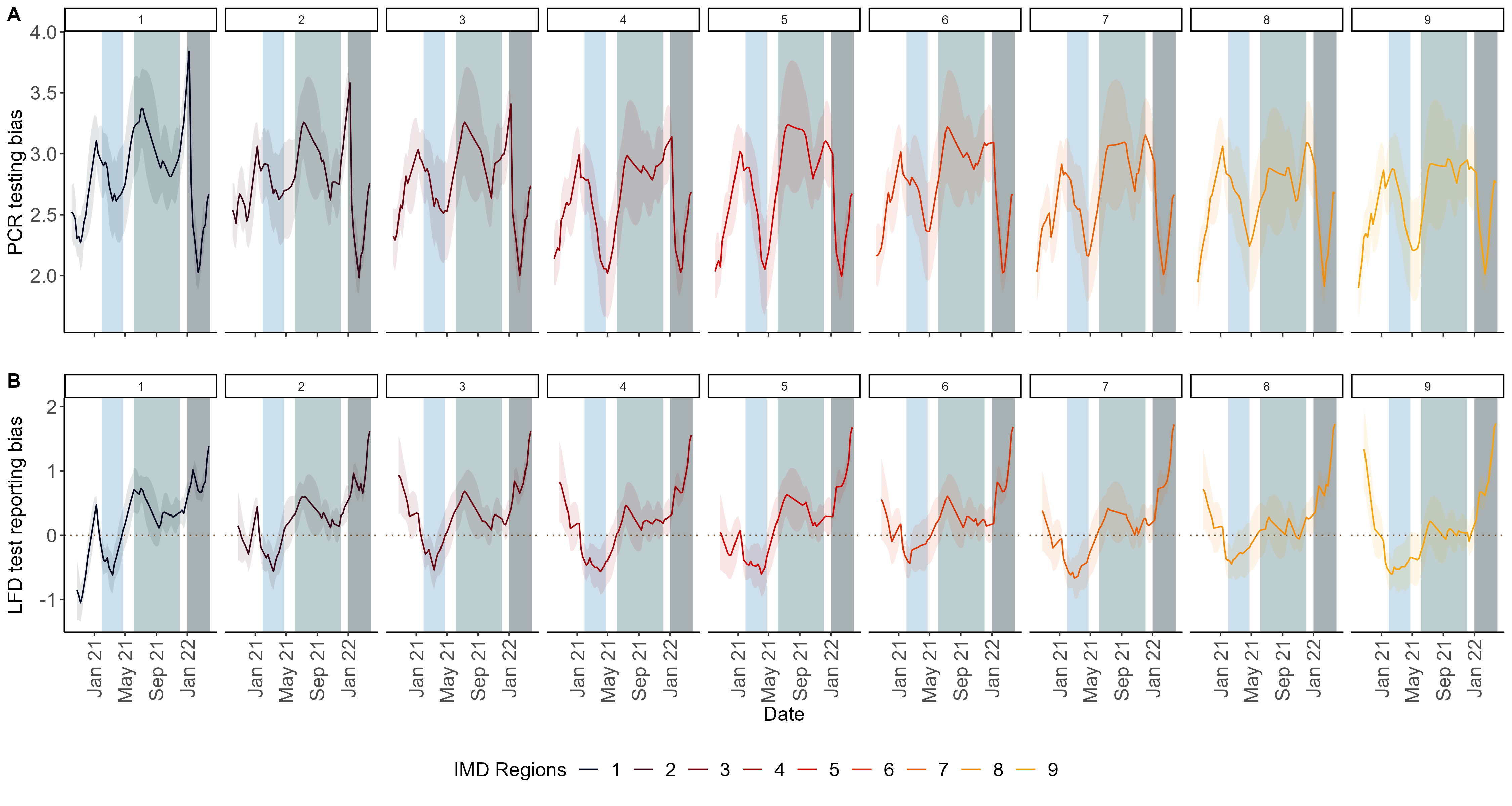

**Figure S13: Testing bias with uncertainty intervals by IMD.** Estimated testing bias defined as the log odds of performing (or reporting) a PCR (or LFD) test in the infected versus uninfected subpopulations by deprivation levels. The solid curves are the posterior means, and the shaded areas are the 95% central credible intervals.

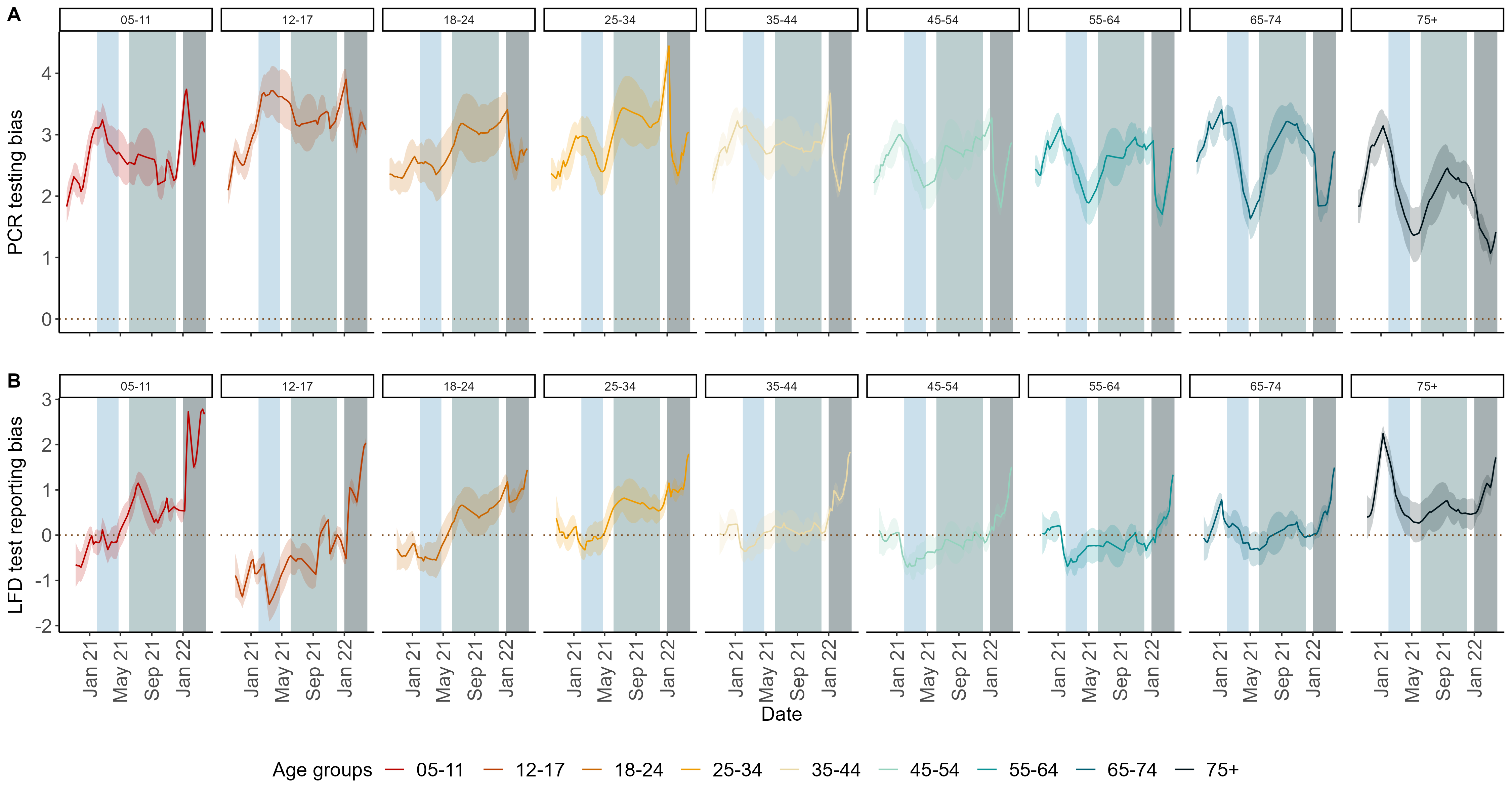

**Figure S14: Testing bias with uncertainty intervals by age-groups.** Estimated testing bias defined as the log odds of performing (or reporting) a PCR (or LFD) test in the infected versus uninfected subpopulations by age groups. The solid curves are the posterior means, and the shaded areas are the 95% central credible intervals.

### Data pre-processing

#### LTLA codes

Due to (Lower Tier Local Authority) LTLA boundary reassignments, some historical LTLA codes are now inactive, so we updated them and assigned regions (as given by the Office of National Statistics) where necessary according to Table S1 below.

**A**

| **Inactive LTLA codes** | **New LTLA codes** |
| --- | --- |
| E07000004, E07000005, E07000006, E07000007 | E06000060 |
| E06000029 | E06000058 |
| E07000049 | E06000059 |
| E07000150, E07000152, E07000153, E07000156 | E06000061 |
| E07000151, E07000154, E07000155 | E06000062 |
| E07000205, E07000206 | E07000244 |
| E07000201, E07000204 | E07000245 |
| E07000190, E07000191 | E07000246 |

**B**

| **LTLA code** | **Region** |
| --- | --- |
| E07000004, E07000005, E07000006, E07000007 | South-East |
| E07000009 | East |
| E07000150, E07000151, E07000152, E07000153, E07000154, E07000155, E07000156 | East Midlands |

**Table S1:** (A) LTLA code reassignment for inactive codes. (B) Regions assigned to LTLAs where this information was missing.

The Isles of Scilly and City of London had no data on ethnicity (LTLA codes = E06000053 and E09000001 respectively) and were removed from the ethnicity analysis.

#### Deprivation

Indices of Deprivation (IoD2019) are based on 39 separate indicators, organised across seven distinct domains of deprivation (Income Deprivation (22.5%), Employment Deprivation (22.5%), Education, Skills and Training Deprivation (13.5%), Health Deprivation and Disability (13.5%), Crime (9.3%), Barriers to Housing and Services (9.3%), Living Environment Deprivation (9.3%)) which are combined and weighted to calculate the Index of Multiple Deprivation 2019 (which we abbreviate as `IMD’). This is an overall measure of multiple deprivation experienced by people living in an area and is calculated for every Lower-layer Super Output Area (LSOA) and was aggregated to other administrative levels by the ONS. The LTLA level measure was calculated by taking the population weighted average of all the LSOA scores in each LTLA. Lastly, each LTLA was assigned deciles according to this aggregated `IMD- Rank of average score`^2^. E06000060, E06000061, E06000062, do not have an IMD assigned to them in the latest dataset and were dropped from the analysis.

#### Ethnicity

The ethnic population data were available only to the nearest thousand^3^. These estimates (published by the Office of National Statistics on 04^th^ December 2019) were based on the Annual Population Survey (which is the Labour Force Survey plus various sample boosts), the mid-year population estimates and 2011 Census, using the method described here^4^. About half (53%) of the LTLAs had zero as inputs for ethnic group populations, which meant that the true number could be a fraction between zero and one (in thousands). In this case, we assumed 0.5 (in thousands) or 500 individuals to belong to such a group in a given LTLA. We now describe the process of assigning ethnic population sizes by LTLAs to match the 2020 population estimates^5^. Let $Y_{ij}^{0}$ be the population (in thousands) of the $i^{th}$ ethnic group in LTLA $j$, and $\sum_{i} Y_{ij}^{0}=Y_{j}^{0}$ was the population of LTLA $j$ from the ethnic population data from 2019^3^. According to the assumption mentioned above, we updated $Y_{ij}^{0}$ as follows:

$$Y_{ij}= \left\{ \begin{aligned} Y_{ij}^{0}, \mathrm{if} Y_{ij}^{0}>0, \\ 0.5, otherwise, \end{aligned} \right.$$

(1)

and $\sum_{i} Y_{ij}=Y_{j}$ was the updated population size of LTLA $j$. We then calculated the proportion $p_{ij}$ of ethnic group $i$ in LTLA $j$ as:

$$p_{ij}= \frac{Y_{ij}}{Y_{j}}$$

(2)

These proportions were then multiplied by the mid 2020 population size estimates^5^ for the LTLAs $Y_{j}^{new}$, to obtain scaled up new ethnic group populations in LTLAs $Y_{ij}^{new}$:

$$Y_{ij}^{new}=p_{ij}Y_{j}^{new}.$$

(3)

In the Pillar 2 mass PCR and LFD testing data, individuals self-reported their ethnicities and had the choice to select the option of “Prefer not to say/Unknown”. We reclassified these responses in the ethnicity column according to the proportion of other ethnicities for each day (or week) in a given LTLA.

Let us consider the case of distributing $X_{un,i,t}$ of say, LFD positive cases reported in the $i^{th}$ LTLA during week $t$. The number (and proportion) of LFD positive cases reported in this LTLA with self-reported ethnicities for each of the $eth$ ethnic groups: “Asian or Asian British”, “Black; African; Black British or Caribbean”, “White”, “Mixed or Multiple ethnic groups”, and “Other ethnic group” was defined as: $X_{eth,i,t} (p_{eth,i,t}=X_{eth,i,t}/X_{i,t})$; where $X_{i,t}= \sum_{eth} X_{eth,i,t}$ was the number of LFD positive cases with self-reported ethnicities in LTLA $i$ during week $t$.

Then $X_{un,i,t}$ was redistributed and the updated numbers for the known ethnic groups was given by:

$$X_{eth,i,t}^{new}= {X_{eth,i,t}+(p}_{eth,i,t}X_{un,i,t}).$$

(4)

Say there were 10 LFD positive cases ($X_{un,i,t}$) reported during a given week in an LTLA that were classified as “Prefer not to say/Unknown” ethnicity, and $X_{eth,i,t}$was disaggregated as the following: say there were $X_{a,i,t} =20$ LFD positives classified as “Asian or Asian British”; $X_{b,i,t}=30$ as “Black; African; Black British or Caribbean”; $X_{w,i,t}=30$ as “White”; $X_{m,i,t}=10$ as “Mixed or Multiple ethnic groups”; and $X_{o,i,t}=10$ as “Other ethnic group”. Then after the redistribution the number would be $X_{a,i,t}^{new}=20+\left( 0.2*10 \right)=22, X_{b,i,t}^{new}=30+\left( 0.3*10 \right)=33, X_{w,i,t}^{new}=30+\left( 0.3*10 \right)=33, X_{m,i,t}^{new}=10+\left( 0.1*10 \right)=11, X_{o,i,t}^{new}=10+\left( 0.1*10 \right)=11$, respectively.

#### Age

Age group bounds were not always consistent between REACT-1, mass testing datasets, and population estimates, and we assumed equivalence between age groups as described in Table S2 below.

| REACT-1 | Testing and population size data |
| --- | --- |
| 5-11 | 4-11 |
| 12-17 | 12-17 |
| 18-24 | 18-23 |
| 25-34 | 24-33 |
| 35-44 | 34-43 |
| 45-54 | 44-53 |
| 55-64 | 54-63 |
| 65-74 | 64-73 |
| 75+ | 74+ |

Table S2: Ages (in years) included in age-group bins in REACT-1 and in either the Pillar 2 testing data obtained from UKSHSA or the population size data.

### Variant time periods

Data

To calculate the weekly frequencies of Variants of Concern (VOCs) in England, i.e., the weekly proportion of SARS-CoV-2 infections that are of a particular VOC, we used weekly lineage counts from genomes sequenced at the Wellcome Sanger Institute, downloaded from <https://covid19.sanger.ac.uk/downloads> (last accessed on 28 July 2023). Lineages are assigned according to the PANGOLIN nomenclature^6^, and the number of each lineage is enumerated by week (according to sample dates) and Lower Tier Local Authority (LTLA).

Method

To obtain estimates of the weekly lineage frequencies at the national level, lineage counts were aggregated over all LTLAs in England. We considered only the major waves of VOC that occurred during the study period, so weekly genome counts were further aggregated according to whether the lineage belongs to Alpha (all sub-lineages of B.1.1.7 and Q), Delta (all sub-lineages of B.1.617.2 and AY), and Omicron (all sub-lineages of B.1.1.529 and BA). Lineages that do not fall under these three VOCs were considered as “wild-types”. The weekly frequencies of these three VOCs and the wild-type of SARS-CoV-2 were calculated at weekly intervals throughout the study period. To estimate the time periods during which these three VOCs were dominant, we used the first and last date when the frequencies of each VOC exceeded [90%]. The time periods where each variant was dominant are as follows:

- Alpha: 30^th^ January 2021 to 24^th^ April 2021
- Delta: 5^th^ June 2021 to 4^th^ December 2021
- Omicron: 1^st^ January 2022 to 14^th^ May 2022 (or end of our study period on 31^st^ March 2022)

### Causal debiasing approach for obtaining time-varying estimates of SARS-CoV-2 prevalence at fine scale resolution and for estimating testing biases

#### Method

Understanding patterns of transmission at fine scales is crucial for determining targeted and effective intervention strategies, particularly because transmission is thought to respond to local conditions. In this section, we apply the method proposed by Nicholson et al.^7^ which combines the mass PCR (or LFD) testing data from Pillar 2 with the REACT-1 prevalence survey data, to produce fine scale estimates of prevalence for sociodemographic groups.

In essence, this method compares REACT-1 and the Pillar 2 mass testing data at a ``coarse’’ level (e.g., administrative regions, IMD regions, or age-group) and outputs a testing bias, which explicitly is given by:

$$omega=\log(\frac{\mathrm{odds}(\mathrm{tested}|\mathrm{infected})}{\mathrm{odds}(\mathrm{tested}|not infected)})$$

(5)

omega at this coarse level is then used to correct Pillar 2 data at a lower level (e.g. LTLA or ethnicity within LTLA or age-group within LTLA) to produce estimates of SARS-CoV-2 prevalence at this scale.

The REACT-1 data consisted of numbers of tests conducted by week within each lower tier local authority (LTLA), along with the results of these tests. In week-LTLAs with fewer than five observations, the data were censored if there were any positives to ensure the data were not identifiable.

Here we describe the coarse and fine levels for the deprivation, ethnicity and age-group analyses using this method and the summary is provided in Table S3.

##### Deprivation

For the deprivation analysis, the aim was to estimate PCR and LFD testing bias (Figure 4A, 4C) for deprivation levels in England. We used the nine IMD levels in England as the coarse level. All LTLAs were assigned an IMD level (1 and 9 corresponding to most deprived and least deprived respectively) as described in the data pre-processing section. Testing data from these LTLAs were aggregated to form nine coarse IMD levels with corresponding REACT-1 and PCR Pillar 2 testing data on the number of tests performed and number of positives. The testing bias (Figure 4A) was estimated for each IMD level and propagated to the LTLAs nested within the coarse levels to produce fine scale prevalence estimates. This assumes that the bias which is influenced by PCR test seeking behaviour, testing capacity and PCR sensitivity and specificity along with other confounding factors, is homogeneous within a given coarse level. We plot these prevalence estimates for a selection of LTLAs along with their corresponding REACT-1 and PCR estimates in Figure SM1. We also plot national deprivation level-specific prevalences by a weighted mean of our fine scale LTLA prevalences and respective population sizes (Figure S6A). Similarly, LFD test reporting bias was also estimated at the coarse IMD levels (Figure 4C).

##### Ethnicity

For ethnicity-related prevalence analysis, the coarse level comprised the nine administrative regions in England, and the five ethnic groups (as described above) in each LTLA made up the finer level of analysis. For example, the ethnic group of Whites in Oxford (LTLA code = E07000178) was nested in the South-East administrative region of England in this analysis. This allowed us to estimate SARS-CoV-2 prevalences for ethnic groups within LTLAs. This implicitly assumed that all the LTLAs and their ethnic groups within a specific administrative region had the same PCR testing bias and behaviour since we did not have access to coarser ethnic group REACT-1 data. For plotting purposes, we obtained national (or regional) ethnic-group prevalences by a weighted mean of our fine scale ethnic-group SARS-CoV-2 prevalence estimates and the respective population sizes of these groups in LTLAs (Figure 2A).

##### Age

For the age-specific analysis, nine age-groups (as described in Table S2) at the national level were used as the coarse level; for the fine scale, we used the LTLA-specific age-groups. We estimated testing biases between age-groups in England (Figure 4B, 4D) and fine scale prevalences for age-groups within LTLAs. For national level plots, we used a weighted mean of the fine scale COVID-19 prevalence estimates and the respective population size of each age-group within an LTLA (Figure 2B).

##### LTLA

While it is not a key output of this paper, we also estimated LTLA-specific prevalences when these fine levels were nested in their nine administrative regions of England (as done previously^7^, Figure SM2 shows the regional model bias estimates). This was used for model comparison as described in the model comparison/sensitivity analysis section below, to plot the spatial prevalence maps (Figure S4), and to calculate national incidence and case detection ratio as well as its association with IMD (see section Incidence and case detection ratio estimation and Figure 5A, 5B).

##### LTLA prevalence and PCR tests

To test if PCR testing quantities were associated with prevalence, we performed weekly $(t)$ simple linear regressions between LTLA $(i)$ specific prevalences $p_{t,i}$ (estimated according to the methodology explained in Deprivation subsection above) and PCR tests performed per capita $n_{pcr,t,i}$ as follows:

$${n_{pcr,t,i}=\beta_{0,t}+\beta_{1,t}p}_{t,i}$$

Then using the observed $p_{t,i}$ and estimated $\beta_{0,t}$ and $\beta_{1,t}$, we predicted the weekly LTLA specific number of PCR tests performed per capita $\hat{n}_{pcr,t,i}$. We then used the weekly medians and interquartile ranges of these predicted values by IMD to plot Figure S6C.

| **Sociodemographic group** | **Fine scale levels** | **Coarse levels** | **Testing bias estimated at coarse level (Figures*)** | **Prevalence estimated at fine scale level (Figures*)** | **Prevalence estimates used in incidence estimation** |
| --- | --- | --- | --- | --- | --- |
| Deprivation (IMD) | LTLAs | National IMD levels | Yes (4A, 4C and S13) | Yes (S7A, S7C) | No |
| Ethnicity | Ethnic groups within LTLAs | Administrative regions of England | No | Yes (2A, S2, S11) | Yes (5C) |
| Age | Age groups within LTLAs | National age groups | Yes (4B, 4D and S14) | Yes (2B, S3) | No |
| NA | LTLAs | Administrative regions of England | No | Yes (S4) | Yes (5A, 5B) |

**Table S3:** Summary of fine scale and coarse level grouping for the different debiasing and incidence analyses. * Figures were part of main manuscript or supplementary results. This does not include figures included in the supplementary methods.

#### Model comparison/sensitivity analysis

##### Deprivation

We compared three different groupings to determine if IMD levels were appropriate as coarse levels. (This was done to compare the robustness of the prevalence estimates from our novel coarse level grouping to the published methodology and results^7^.) The IMD-grouped model was a moderately better fit to the data compared to using administrative regions as the coarse level (Table S4). We determined the model’s fit by summing the log-likelihood for the weekly fine scale LTLA prevalences according to the likelihood given by the Binomial distribution as

$$X_{pos,i,t}\sim\mathrm{Binomial}\left( X_{total,i,t}, {prev}_{i,t} \right),$$

(6)

where $X_{total,i,t}$ is the observed count of PCR tests and $X_{pos,i,t}$ is the count of PCR positives in the the REACT-1 survey during week $t$ in LTLA $i$, and ${prev}_{i,t}$ is the posterior mean of the fine scale prevalence estimates. The model with higher likelihood (or log likelihood) is a better fit to the data, i.e. its prevalence estimates for LTLAs over the study period best explain the likelihood of observing the LTLA specific REACT-1 PCR positive data.

We also used a coarse grouping based on both IMD and administrative regions because both groupings in isolation gave similar model fits (Table S4) and we hypothesised that together they might be an even better fit to the data; in doing so we used only five deprivation levels (instead of nine like above) to ensure there were enough LTLAs in each category, resulting in 41 coarse groups overall. This model had the highest predictive accuracy (Table S4). In this grouping, the trends in PCR testing bias were more idiosyncratic, although there were regions and time periods where the bias was higher in more deprived areas. For example, during week of 1^st^ June 2021 (Figure SM3) the estimated odds of testing were exp(3.17) ≈ 23 and exp(1.89) ≈ 6 times higher in individuals with infection compared to individuals without infection in most deprived (IMD = 1) and least deprived areas (IMD = 5) in North West England.

| **Coarse level** | **Log-likelihood** |
| --- | --- |
| 5 IMD regions and 9 regions | -106,594 |
| 9 IMD regions | -107,138 |
| 9 Regions | -107,330 |

**Table S4:** **Model comparison results across different coarse groupings.** The models were fit at the coarse level (administrative region, IMD or administrative region and IMD) meaning that the predictions made at the LTLA level were largely independent of LTLA specific characteristics (apart from the fact they belong to the larger group). However, as the size of coarse groups gets smaller, there is greater correspondence between the LTLA level characteristics and those of the coarse groups to which they belong. Because of this, our estimate of predictive accuracy between the model fit to 9 administrative regions and 9 IMD regions is likely to be more comparable than the model fit to 41 regions.

We were unable to do similar model comparisons for ethnicity and age-group prevalence analysis because we did not have access to fine scale data on the number of tests and positives by ethnic group or age group within LTLAs from REACT-1.

##### Ethnicity

For LTLAs with zero (in thousands) population for a given ethnic group, we performed sensitivity analyses by considering 0.2 as well as 0.8 thousand as population sizes in the two LTLAs with highest and lower proportion of Whites each and saw similar prevalence estimates (see Figure SM4).

### Incidence and case detection ratio estimation

SARS-CoV-2 prevalence (we defined an individual to be infection-positive if their infection was detectable by PCR with cycle threshold (Ct) value < 37) is dependent on recent histories of infection occurrence, which we model using the following equation (with both SARS-CoV-2 prevalence and infection occurrence measured on the same scale):

$\theta_{t}=\sum_{\tau=1}^{\infty} \omega_{\tau}i_{t-\tau}$,

where $\theta_{t}$ denotes prevalence on day $t$ and $i_{t}$ is the incidence on the same day; $0 \leq\omega_{\tau} \leq1$ is the probability of being PCR positive given an infection which began $\tau$ days ago.

If the set of probabilities denoting PCR positivity $\left\{ \omega_{\tau} \right\}_{\tau=1}^{\infty}$ were precisely known, the above equation provides a linear system which can be inverted to determine an incidence time series $\left\{ i_{t} \right\}_{1}^{T}$. But these probabilities are typically determined from regular testing data on individuals whose date of exposure is known or can be determined with a degree of certainty^8^, and these probabilities likely changed throughout the course of the pandemic with the introduction of novel SARS-CoV-2 variants^9^. To account for these imperfections in our knowledge of $\left\{ \omega_{\tau} \right\}_{\tau=1}^{\infty}$,, we assumed a probabilistic relationship between incidence and prevalence of the form:

$\theta_{t}\sim N\left( \sum_{\tau=1}^{\infty} \omega_{\tau}i_{t-\tau},\sigma\right)$,

where $N\left( \mu,\sigma\right)$ represents a normal distribution with mean $\mu$ and standard deviation $\sigma$; we assumed that the $\theta_{t}$ were independent random variables, which would likely be violated in presence of substantial autocorrelation in the misspecification of $\left\{ \omega_{\tau} \right\}_{\tau=1}^{\infty}$; $\sigma$ is estimated and characterises the degree of imperfections in our model.

We used a Bayesian framework to estimate $\left\{ i_{t} \right\}_{1}^{T}$ and assumed a prior as described in the main text, which assumed that the true infections were a constant to-be-estimated multiple of the reported cases that was assumed piecewise-constant with pieces of length 15 days. Since the reported cases are the dates the cases appear in the testing data, the date of infections likely precedes these, and our assumption is that this lag is minimal relative to longer-term changes in case reporting behaviours.

Our approach to estimate the true case incidence as described above required daily estimates of prevalence and how the PCR detectability varied over the course of a typical infection throughout the study period. We also chose to use positive test results reported via UKHSA’s Pillar 2 PCR and LFD data to improve the precision of our incidence estimates. We elaborate on these three inputs below.

#### Daily SARS-CoV-2 prevalence estimates

For Figure 5A we used LTLA-specific debiased prevalence estimates obtained using the methodology explained in the section “Causal debiasing approach for obtaining time-varying estimates of SARS-CoV-2 prevalence at fine scale resolution and for estimating testing biases”, where the LTLAs were nested within the nine administrative regions of England. These were used to produce LTLA-specific incidences and then aggregated to produce national incidence estimates. We also used publicly available national prevalence estimates published by ONS^10^ to obtain another set of national incidence estimates.

For Figure 5B we estimated incidence for each LTLA by using prevalence obtained from our debiasing approach, when we assumed the LTLAs were the fine scale regions, and the administrative regions were the coarse scale regions.

For the ethnicity analysis, we used the mean prevalence estimates for each ethnic group within LTLAs of England.

A summary of the prevalences used and associated figures in the main manuscript are provided in Table S3.

For the three prevalence time series described above, the SARS-CoV-2 prevalence was itself estimated with a margin of uncertainty, and we included this uncertainty in our analysis via a prior probability distribution which links the true prevalence with that estimated by our incidence model (<https://github.com/sumalibajaj/equitytesting>). The debiasing approach^7^ estimated mean prevalence and with uncertainty bounds, and the prevalence estimates from ONS had upper and lower uncertainty bounds. We assumed that this uncertainty was captured by a normal distribution,

$$\theta_{t}\sim\mathrm{normal}\left( \theta_{t,central}, \sigma_{\theta,t} \right),$$

(7)

where $\theta_{t}$ was the prevalence on day $t$ as described in the main manuscript and $\theta_{t,central}$was the mean prevalence estimate from the debiasing methodology or as published by ONS, and $\sigma_{\theta,t}$ was the uncertainty in these estimates. Standard errors were back-calculated from each of the lower ($\theta_{t,lower}$) and upper ($\theta_{t,upper}$) bounds for weekly prevalences of each fine scale level and national ONS estimates as follows:

$$\sigma_{\theta,t,upper}=\frac{\theta_{t,upper}-\theta_{t,central}}{1.96}$$

$$\sigma_{\theta,t,lower}=\frac{\theta_{t,central}-\theta_{t,lower}}{1.96}$$

(8)

we then took the average of these two standard errors and used it as the standard deviation of the specified normal priors for prevalence in our model:

$$\sigma_{\theta,t}= \frac{\sigma_{\theta,t,upper}+\sigma_{\theta,t,lower}}{2}.$$

(9)

Since the models we used were formulated at the daily level, we approximated daily prevalence (and its standard error) by linearly interpolating the weekly measures. This likely understates the uncertainty in daily prevalence but this source of uncertainty was unlikely to have affected the qualitative trends we found across longer timescales.

#### The probability of detectable infection by PCR as a function of time after infection

For the pre-Omicron period (defined as the period before 31^st^ Dec 2021 in our analysis), we extracted the posterior median probability of PCR detectability for each day subsequent to infection up to a maximum of 30 days (after which the probability of testing positive via PCR was estimated as approximately zero^8^) from Hellewell et al.^8^ (Figure SM5). We assumed that the PCR detectability time curve was the same for all time periods before Omicron; but since there was evidence that this changed for Omicron, we used updated estimates^9^ starting from 1^st^ Jan 2022 to end of our study period. We used the frequent testing data provided by Hay et al.^9^ and estimated the proportion of individuals with detectable infection by SARS-CoV-2 Omicron VOC (PCR CtT1 value<37) for 30 days since detection (Figure SM5). Since these Omicron estimates represented time since detection opposed to infection, we crudely shifted the Omicron estimates by one day forward and used Hellewell et al.’s^8^ day one estimate of PCR detectability. Figure SM5 provides a comparison between the two profiles of PCR detectability up until 30 days since infection. The method required at least 30 days of prevalence data to estimate incidence because we assumed that the probability of detecting infection via PCR after 30 days since infection was zero^8^.

#### Positive SARS-CoV-2 mass testing data

Since the reported PCR and LFD positive data had weekly fluctuations, likely due to reporting biases on certain days of the week, we smoothed the daily reported cases data using a 7-day moving average centred on the day under consideration.

#### Model fitting

Because of the computational expense of full uncertainty quantification via Markov chain Monte Carlo (MCMC), we used optimisation to fit our model to data, resulting in a single set of parameters characterising the *maximum a posteriori* (MAP) estimates. The model was specified using the Stan language and fitted via their default optimisation algorithm^11^ with the highest log-probability estimates amongst 5 separate optimisations constituting our parameter estimates. Figure SM6 compares the MAP and MCMC estimates for a subset of LTLAs with the highest and lowest proportion of individuals in the White ethnic groups, and shows that for these, the estimated uncertainty obtained using MCMC was minimal.

#### Model fit checks

As discussed above, we produced two sets of incidence estimates, one for each source of prevalence data: the prevalence estimates outputted from our debiasing methodology and another set from ONS. We were able to check the fit of our model to the data by comparing the prevalence estimates determined by our model with those prevalences used to fit it. For the ONS prevalence model, this was straightforward to visualise, and the fit to the data was reasonable (see Figure SM7). For the LTLA-level prevalence model fits, we computed the root-mean-square error (RMSE) in predictions (using the central prevalence estimates from the debiased Pillar 2 prevalences). In Figure SM8, we overlay our model-estimated prevalences with the LTLA-level estimates used to fit our model for the four worst- and four best-fitting LTLAs. This showed that, generally, our modelled estimates were a reasonable fit to the data, although there was a general reduction in model performance during the Omicron period – either due to issues with the prevalence estimates used to fit the model or due to inappropriate estimates of the probability of detecting infection by PCR throughout the course of infection.

### Estimating false positivity rate, LFD test sensitivity and specificity

We now describe our approach for estimating group-specific test sensitivities and specificities. To do so, we collated a dataset within our Pillar 2 mass testing data of all reported positive LFD results which were followed up with a PCR test within 3 days of the reported positive. Using these data, we computed the probability of positive confirmatory PCR test within 3 days of reporting a positive LFD test as follows.

For ease of notation, let $\delta$ indicate “reporting a positive LFD test” and $\gamma$ indicate “seeking a PCR test within 3 days of reporting an LFD positive test”. Then using Bayes’ rule,

$$P\left( PCR+ | LFD+,\delta, \gamma\right)=\frac{P\left( LFD+ | PCR+,\delta,\gamma\right) P\left( PCR+ | \delta,\gamma\right)}{P\left( LFD+|\delta,\gamma\right)}$$

(10)

By making the simplifying assumption that $\delta$ and $\gamma$ do not influence the computed probabilities, we can then write the denominator as:

$$P\left( LFD+ | PCR+ \right) P\left( PCR+ \right) +P\left( LFD+ | PCR- \right) P(PCR-)$$

Assuming that PCR tests are perfect, we write:

$P\left( PCR+ | LFD+ \right)=Y$,

$P\left( PCR+ \right)=\mathrm{prevalence}=X$ ,

$P\left( LFD+ | PCR+ \right)=\mathrm{sensitivity}=a$,

$$P\left( LFD+ | PCR- \right)=1-\mathrm{specificity}=b.$$

We can then rewrite (10) as:

$$Y= \frac{aX}{aX+b(1-X)}=\frac{aX}{b+(a-b)X}.$$

(11)

To fit this model to our data, we let number of positive PCR confirmatory tests during week $t$, $n_{pcr+,t},$be follow a binomial distribution:

$$n_{pcr+,t}\sim\mathrm{binomial}(n_{lfd+,t}, Y_{t})$$

(12)

where $n_{lfd+,t}$ is the number of positive LFD tests reported during week $t$ that were followed up with a confirmatory PCR test within 3 days, with $Y_{t}$ modelled according to (11):

$$Y_{t}=\frac{a X_{t}}{b+\left( a-b \right)X_{t}}.$$

(13)

We fit this model to national prevalence data at the group level. where $X_{t}$ is the SARS-CoV-2 prevalence for week $t$, with priors centred around national mean estimates $\mu_{t}$ and standard deviation $\sigma_{t}$. $\mu_{t}$ was obtained as a weighted mean of our fine scales LTLA prevalences and respective population sizes and $\sigma_{t}$ were back-calculated from the corresponding weighted national 95% central credible intervals according to ((8)-(10)):

$$X_{t}\sim\mathrm{normal}(\mu_{t},\sigma_{t}).$$

(14)

The sensitivity and specificity parameters were estimated by fitting the model. These parameters were assigned the following priors:

$$a \sim\mathrm{uniform}\left( 0,1 \right),$$

$$b \sim\mathrm{uniform}\left( 0,1 \right).$$

(15)

We let the sensitivity and specificity vary by ethnic group and age-group. Table S5 describes the estimates of sensitivity ($a$) and 1-specificity ($b$) in models with and without ethnicity or age-group.

It is crucial to note that in addition to LFD tests being reported by individuals themselves, there might be variation in how individuals performed tests, and there may be differences in laboratories and test manufacturers.

| **Model** | **Sensitivity** | **False positivity rate**  **(1- Specificity)** |
| --- | --- | --- |
| No age | 0.68 (0.19, 0.99) | 0.0011 (0.0003, 0.0017) |
| With age (in years) |  |  |
| 5-11 | 0.67 (0.16, 0.99) | 0.0017 (0.0004, 0.0025) |
| 12-17 | 0.67 (0.16, 0.99) | 0.0019 (0.0005, 0.0028) |
| 18-24 | 0.67 (0.16, 0.99) | 0.0013 (0.0003, 0.0019) |
| 25-34 | 0.67 (0.16, 0.99) | 0.0015 (0.0004, 0.0022) |
| 35-44 | 0.67 (0.16, 0.99) | 0.0012 (0.0003, 0.0018) |
| 45-54 | 0.67 (0.16, 0.98) | 0.0011 (0.0003, 0.0016) |
| 55-64 | 0.67 (0.18, 0.99) | 0.0008 (0.0002, 0.0012) |
| 65-74 | 0.67 (0.16, 0.99) | 0.0005 (0.0001, 0.0008) |
| 75+ | 0.67 (0.15, 0.99) | 0.0005 (0.0001, 0.0007) |
| No ethnicity | 0.67 (0.20, 0.98) | 0.0009 (0.0003, 0.0014) |
| With ethnicity |  |  |
| Asian or Asian British | 0.67 (0.18, 0.99) | 0.0011 (0.0003, 0.0016) |
| Black; African; Black British or Caribbean | 0.68 (0.18, 0.99) | 0.0010 (0.0003, 0.0015) |
| Mixed or Multiple ethnic groups | 0.66 (0.16, 0.99) | 0.0016 (0.0004, 0.0024) |
| Other ethnic group | 0.66 (0.16, 0.98) | 0.0007 (0.0002, 0.0011) |
| White | 0.72 (0.32, 0.99) | 0.0009 (0.0004, 0.0013) |

**Table S5:** Posterior median (95% central credible interval) estimates of sensitivity and false positivity rate (= 1-specificity) in models without socio-demographic groups and with ethnic groups and age-groups. Since the number of datapoints in the age-group analysis and ethnicity analysis is not the same, as described in Table 1, we produce two estimates for the model without sociodemographic variables (No age, no ethnicity).

### Modelling the decline in PCR detection rate subsequent to reporting an LFD positive

We modelled how the probability of detecting SARS-CoV-2 infection via PCR after reporting a positive LFD result changed (by assumption, decayed) as a function of time and tested if this rate of decay was different between ethnic groups. Since viral loads are likely modal from the time of infection onwards, it is possible that an individual’s probability of testing positive by PCR could actually increase subsequent to reporting of the LFD positive. The overwhelming trends in our data, however, show that, on average, PCR detectability declined subsequent to the date of reporting of an LFD positive.

To calculate the decay rates, we aggregated two quantities by month $i$, ethnic group $eth$, and by number of days $t$ (where $t=0,1,2,3$) between reporting a positive LFD test and subsequently seeking a PCR test:

i) $n_{lfd+,t,i,eth}$ = number of reported LFD positives that were followed up with a PCR test within $t$ days for ethnic group *eth*,
ii) $n_{pcr+,t,i,eth}$ = corresponding PCR-positive test count.

These were modelled as samples from a binomial distribution:

$$n_{pcr+,t,i,eth}\sim\mathrm{binomial}\left( n_{lfd+,t,i,eth}, p_{t,i,eth} \right),$$

(16)

where $0\leq p_{t,i,eth}\leq1$ was the to-be-estimated probability of PCR detection $t$ days since of reporting a positive LFD result during month $i$ and for a given ethnic group $eth$. This probability was assumed to decay over time according to the following relationship:

$$p_{t,i,eth}=P\left( PCR+ | LFD+,\delta,\gamma,t,i,eth \right)=\varphi_{i,eth}e^{-\lambda_{i,eth}t}.$$

(17)

$\varphi_{i,eth}$ represents the differences in false positivity rates due to monthly changes in prevalences by ethnic groups.

We plot these monthly decay rates by ethnic groups $\lambda_{i,eth}$ in Figure 3A.

To check if our model was a good fit to the data, we plotted the observed and predicted number of $n_{pcr+,t,i}$ or “true positives”. For predicted estimates, we extracted the posterior median of $p_{t,i,eth}$ and multiplied it by $n_{lfd+,t,i,eth}$. Figure SM9 shows that there was agreement between observed and predicted number of confirmatory PCRs.

### Figures for methods

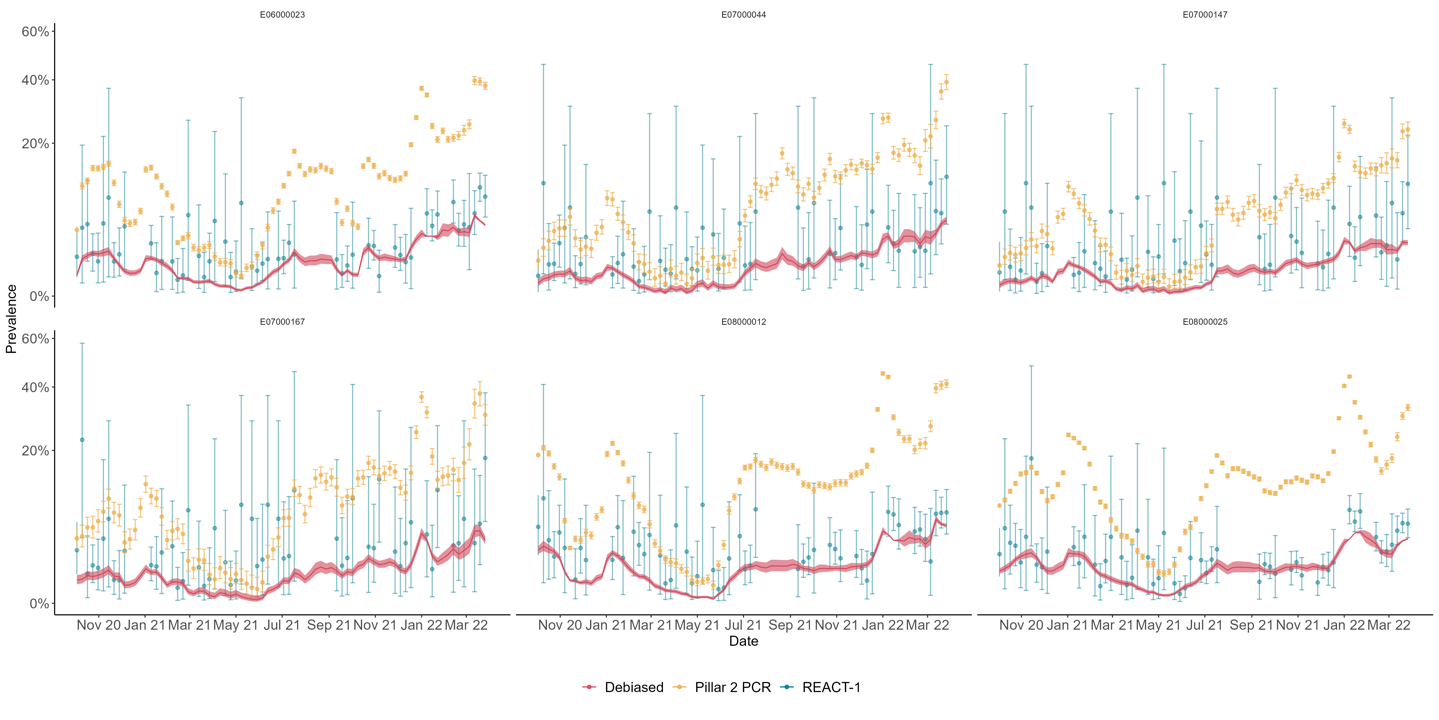

**Figure SM1:** Weekly prevalence estimates for the six LTLAs with highest (first row) and lowest (second row) average weekly prevalence. The colours correspond to different prevalences estimated using data from REACT-1 (blue), Pillar 2 mass testing PCR tests (yellow) and the debiasing method (red). The REACT-1 and Pillar 2 PCR prevalences are plotted with 95% central credible intervals, assuming uniform priors over the probability of testing positive. The shaded area around the debiased prevalence is the 95% central credible interval estimated by the model.

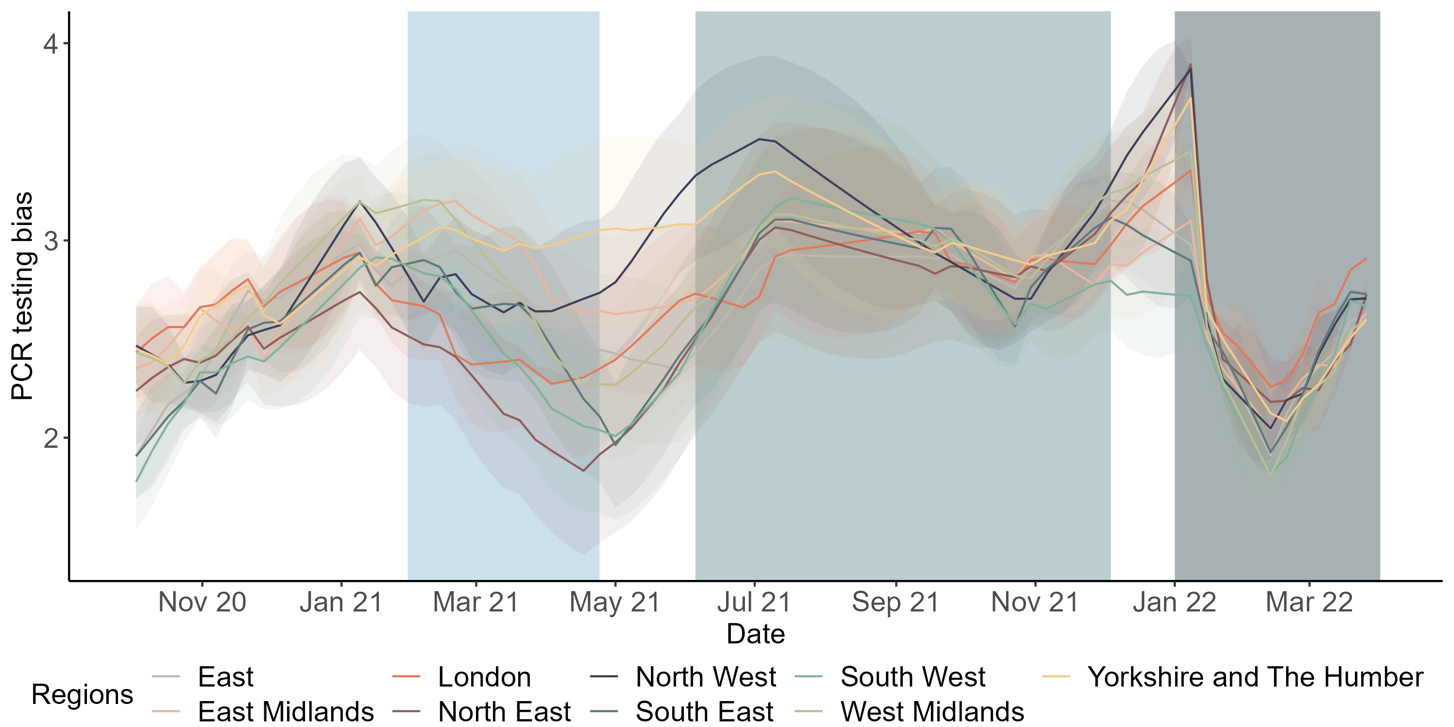

**Figure SM2:** Estimated mean PCR testing bias. I.e. the log-odds of performing a PCR test in the infected versus uninfected subpopulation by administrative regions of England over time and their 95% central credible intervals. The shaded rectangles denote the Alpha, Delta and Omicron time periods.

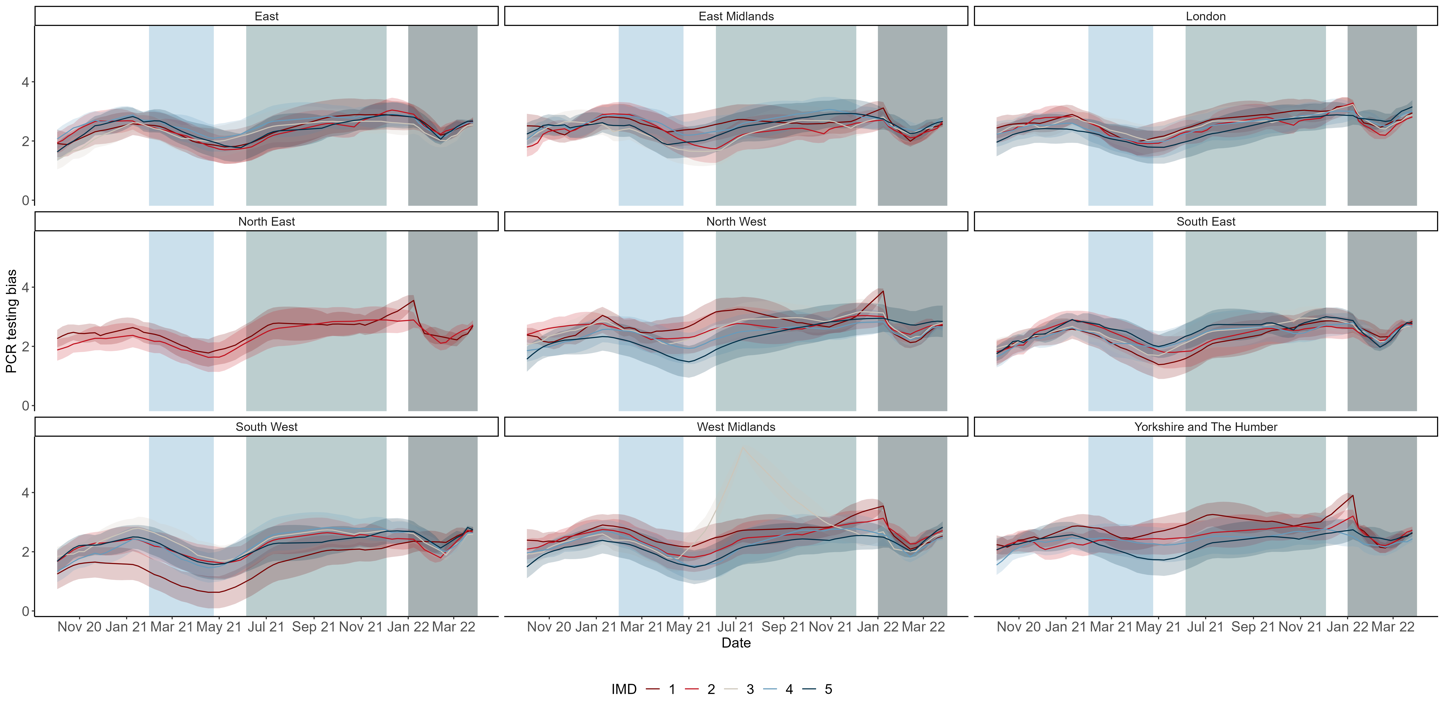

**Figure SM3:** Estimated mean PCR testing bias. I.e. the log-odds of performing a PCR test in the infected versus uninfected subpopulation by deprivation level (IMD 1 = most deprived, 5 = least deprived) and administrative regions of England over time and their 95% central credible intervals.

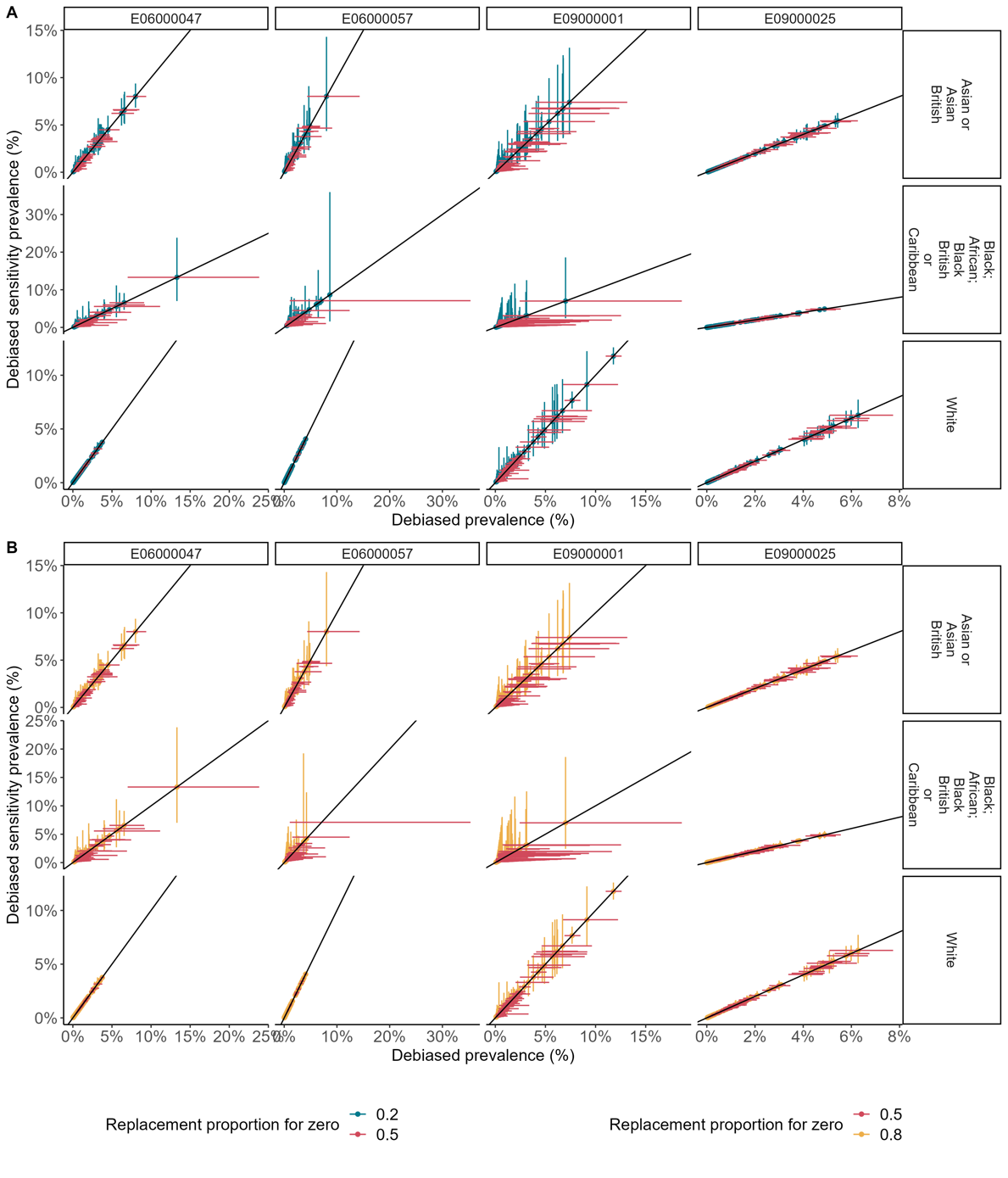

**Figure SM4:** Comparison between ethnic group prevalences for four LTLAs (with first two LTLAs having the maximum proportion of individuals belonging to the White ethnic group; the last two having the minimum) when we replaced the zero proportion of ethnic groups in the data by 0.5 (red and is used in the main ethnic group analysis) by 0.2 (A: blue) and 0.8 (B: yellow). The horizontal and vertical bars are the 95% central credible intervals from the debiasing methodology. The solid 45-degree line is the line to compare how similar the estimates are.

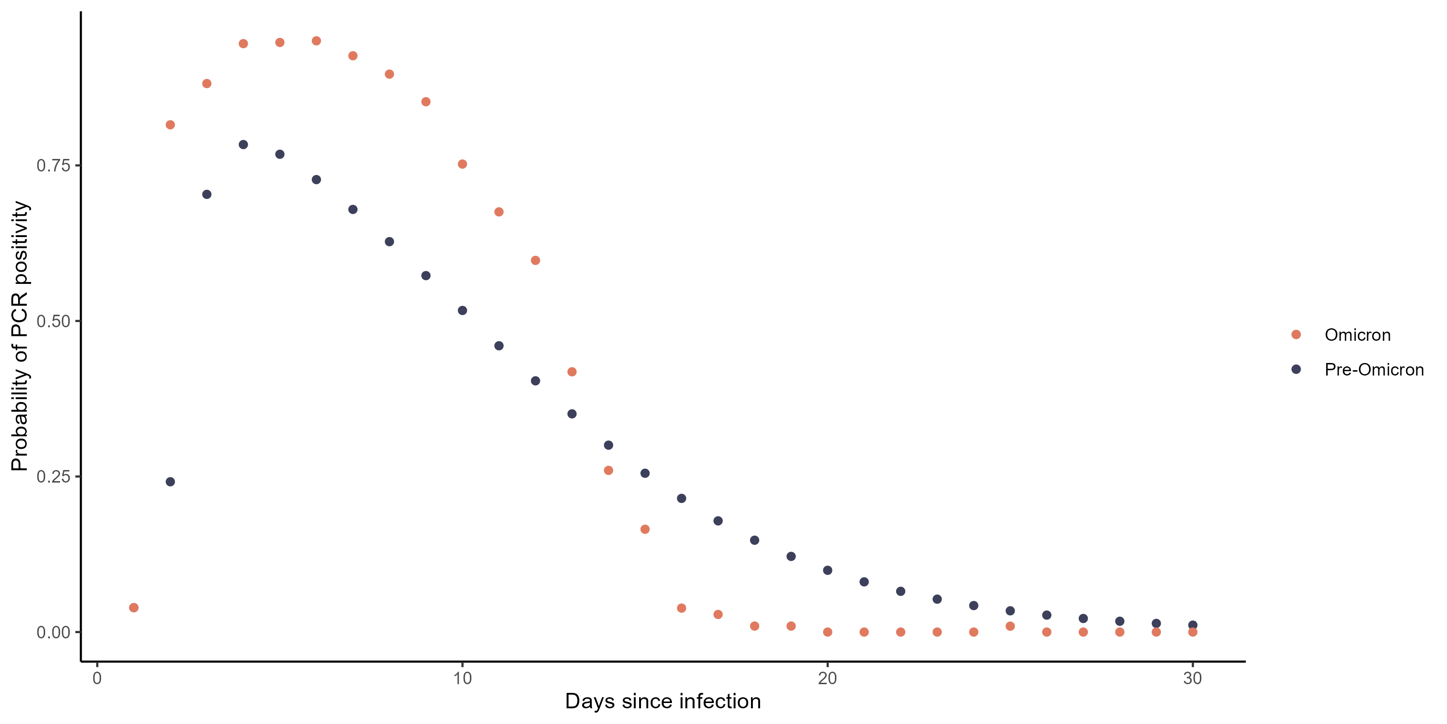

**Figure SM5:** The temporal profile of PCR detectability as a function of time since infection. The dark blue and orange points represent probabilities before and after 15^th^ December 2021, respectively. PCR positivity is defined as Ct value of less than 37.

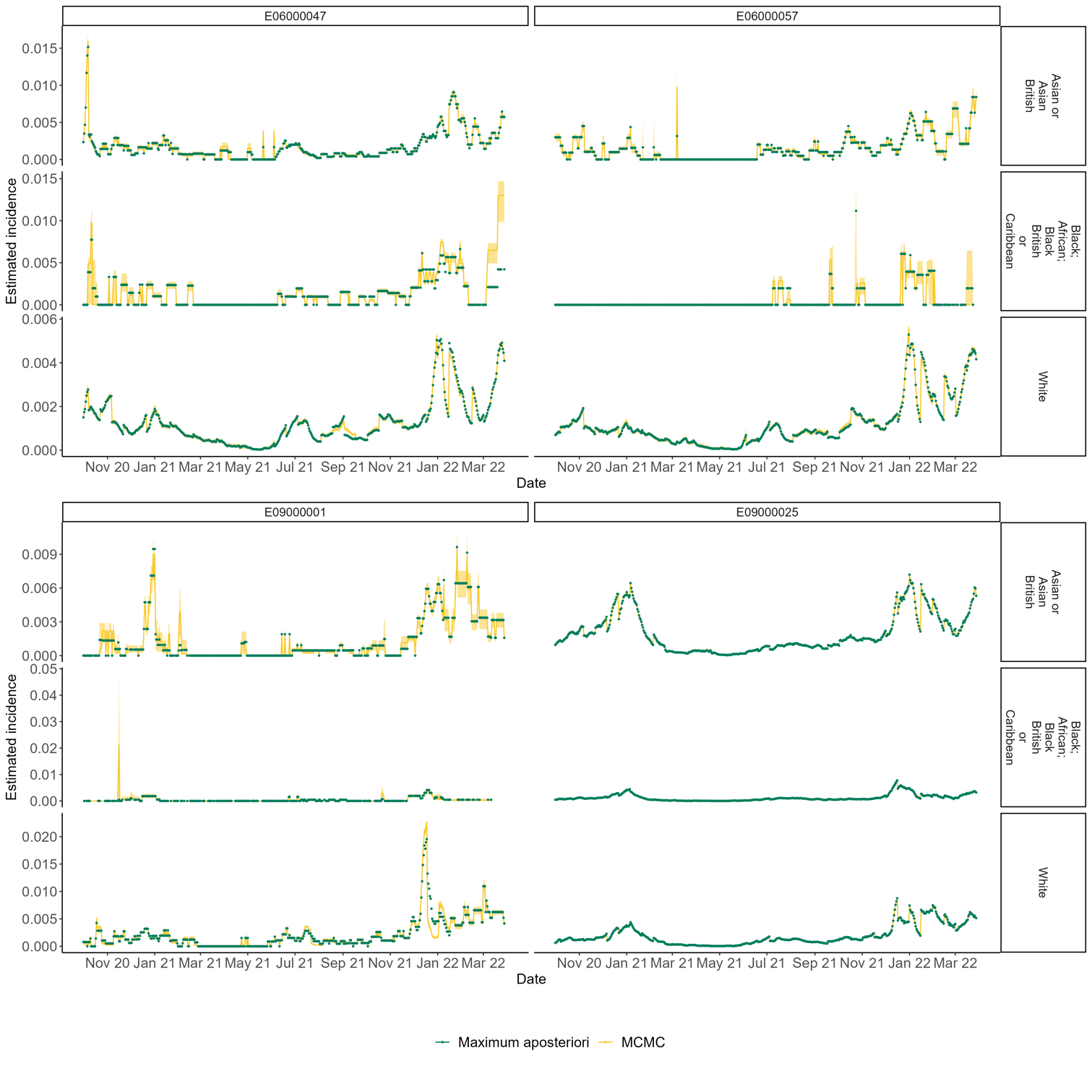

**Figure SM6:** Comparison between the estimated incidence using MCMC and MAP for two LTLAs each with (A) maximum and (B) proportion of individuals in the White ethnic group. The yellow solid curves are the posterior median estimates and the shaded yellow region is the 95% credible interval from the MCMC. The green dots are the parameter estimates with the highest log-probability across five optimisations using Stan’s default optimisation algorithm.

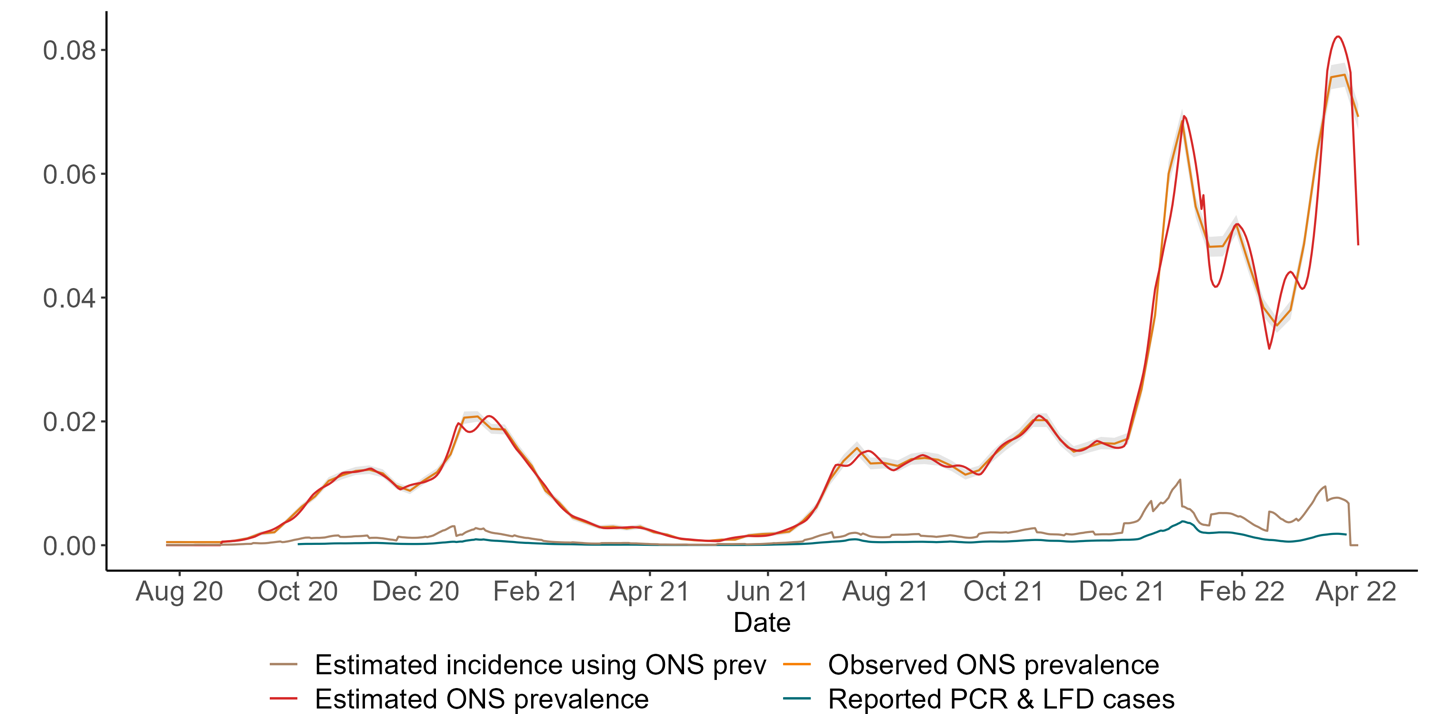

**Figure SM7:** Model fit for the ONS national prevalence model. Comparing country wide estimated (red curve) and observed (solid orange curve and shaded region for 95% central confidence intervals) prevalence from ONS data. The blue and brown curves are the Pillar 2 PCR and LFD positive tests and estimated incidence, respectively, both as a proportion of the population.

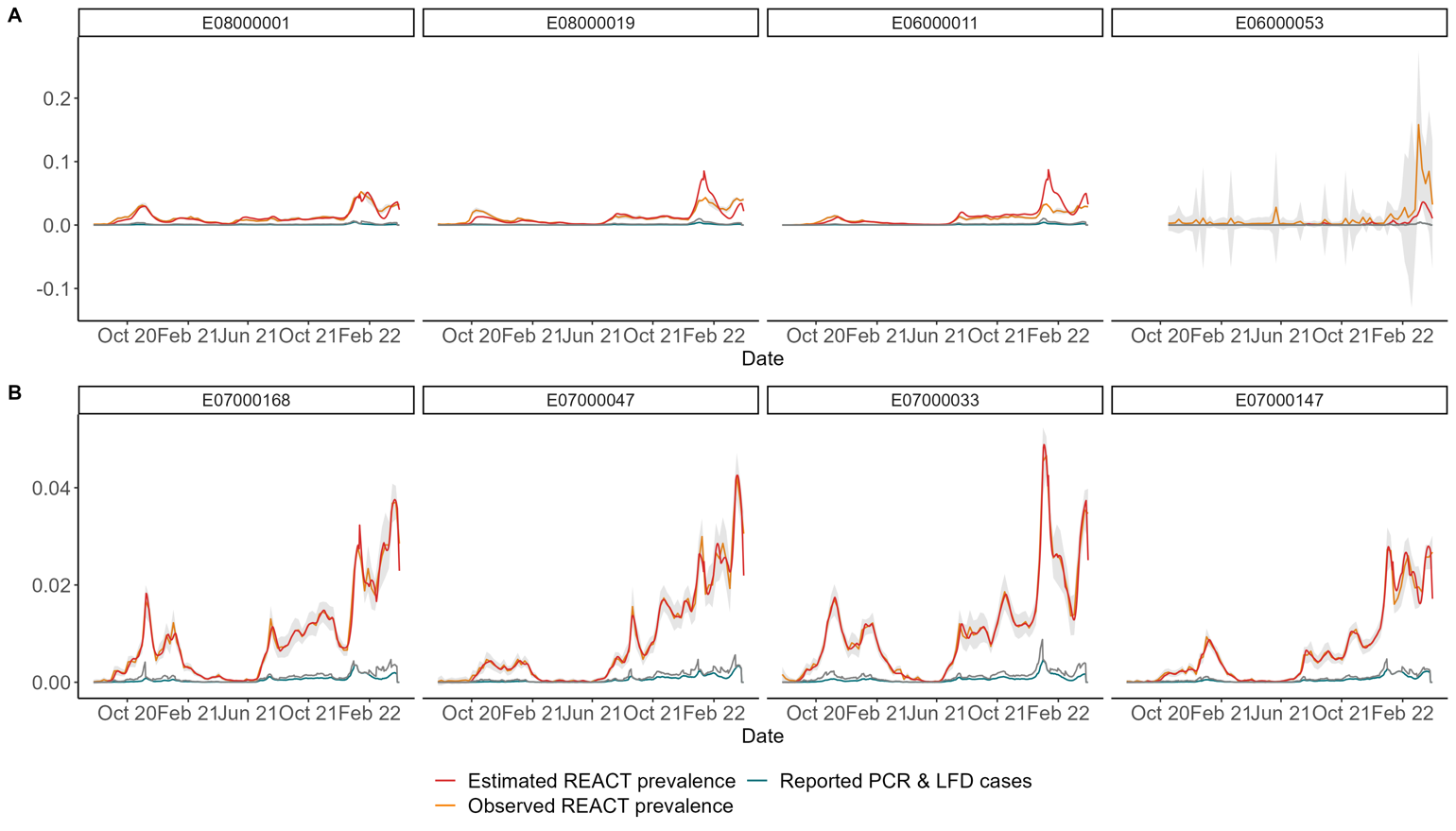
**Figure SM8:** Model fits for the REACT-1 based prevalence model at LTLA level. Comparing countrywide estimated (red curve) and observed (solid orange curve and shaded region for 95% central confidence intervals) prevalence for the (A) four worst and (B) best performing LTLAs, with the highest and lowest RMSEs respectively. The blue and brown curves are the Pillar 2 PCR and LFD positive tests and estimated incidence, respectively, both as a proportion of the population.

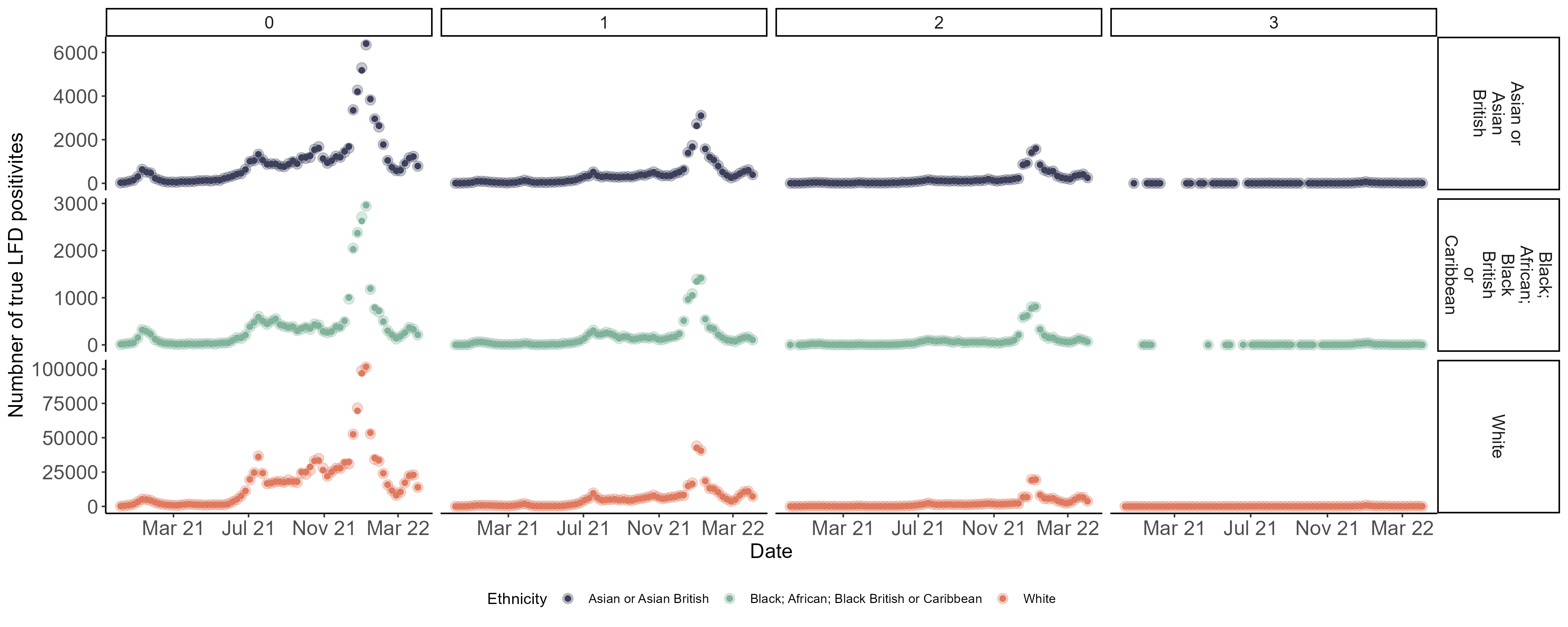

**Figure SM9:** Posterior predictive check for modelling decay rate of PCR detectability after reporting a positive LFD test, by different ethnic groups. The lighter circles are the observed number of “true positives” i.e. positive PCR result within 3 days of reporting a positive LFD. Darker smaller circles are the estimates, for which we extracted the posterior median of probability of PCR positivity and multiplied it by the number of positive LFD reported.

10.	Office of National Statistics. Coronavirus (COVID-19) Infection Survey. https://www.ons.gov.uk/surveys/informationforhouseholdsandindividuals/householdandindividualsurveys/covid19infectionsurvey.

11.	Stan Development Team. Stan Modeling Language Users Guide and Reference Manual, v2.26.1.
